# Restoring intended speech in people with dysarthria from pontine stroke using an intracortical brain-computer interface

**DOI:** 10.64898/2026.02.19.26346583

**Authors:** Samuel R. Nason-Tomaszewski, Pranav I. Deevi, Justin J. Jude, Qinwan Rabbani, Brandon G. Jacques, Anna L. Pritchard, Lahiru N. Wimalasena, Brice A. Richards, Brianna M. Karpowicz, Payton H. Bechefsky, Nicholas S. Card, Angelique C. Paulk, Jian Li, Darrel R. Deo, Eun Young Choi, Leigh R. Hochberg, Sergey D. Stavisky, David M. Brandman, Daniel B. Rubin, Nicholas AuYong, Chethan Pandarinath

**Author notes:** These authors contributed equally (Samuel R. Nason-Tomaszewski, Pranav I. Deevi). These authors contributed equally (Sergey D. Stavisky, David M. Brandman). These authors contributed equally (Nicholas AuYong, Chethan Pandarinath).

## Abstract

Restoring communication for people with dysarthria secondary to pontine stroke remains a critical challenge. Intracortical brain-computer interfaces (iBCIs) have demonstrated great potential for speech restoration in people with amyotrophic lateral sclerosis (ALS), with 1-24% word error rates (WERs) on a 125,000-word vocabulary. In pontine stroke, only BCIs spanning large regions of cortex (using electrocorticography) have been previously tested, achieving 25.5% WERs with a smaller 1,024-word vocabulary. Whether intracortical BCI performance improvements extend to people with pontine stroke-induced dysarthria remains unclear. Here, we detail targeting and deployment strategies for iBCIs that restore intended speech in people with long-standing dysarthria and tetraplegia due to pontine stroke, and show that neural activity from as few as one 64-channel microelectrode array in orofacial motor cortex can achieve high-performance speech decoding, with performance comparable to prior iBCI work. We used neural networks to predict word sequences from iBCI signals as BrainGate2 participants T16 and T22 mimed (mouthed without vocalization) sentences from a large vocabulary. With standard adjustments for signal changes, decoding has remained stable more than two years post-implantation for T16 and two months for T22, achieving median 19.4% and 10.6% WERs with a 125,000-word vocabulary and median 10.0% and 2.25% WERs with a 1,024-word vocabulary for T16 and T22, respectively. Using a more structured task, we demonstrate the day-over-day signal changes that necessitate adjustment. These results demonstrate that brain-to-text iBCIs decoding from small patches of cortex can achieve high performance in individuals with pontine stroke, with comparable performance to speech iBCIs in individuals with ALS.

## Introduction

Communication is often a challenge for people who have had a brainstem stroke, with dysarthria present in 31-49% of cases ^1,2^. People with brainstem stroke causing both tetraplegia and severe dysarthria generally rely on eye-gaze or sip-and-puff systems to communicate; these systems can be fatiguing, unintuitive, and are far slower than conversational communication ^3–6^. Despite the motor impairments that impede communication in brainstem stroke, cortical language networks may remain largely preserved, in contrast to cortical strokes that result in aphasia due to the loss of a critical language network component. This raises the possibility that language-related signals might be extracted from intact cortical regions in people with brainstem stroke, and that those signals could be leveraged to enable more naturalistic, high-throughput communication ^7,8^.

Brain-computer interfaces (BCIs) harness brain activity to restore function. BCIs have enabled people with a variety of forms of paralysis to control computers and prosthetic arms, and more recently there has been tremendous progress developing speech prostheses that predict what words a person is attempting to say, often referred to as a brain-to-text BCI ^9–15^. Specifically for brainstem stroke, electrocorticographic (ECoG) brain-to-text BCIs have decoded surface potentials from ventral sensorimotor brain regions into predicted text, restoring communication to a person with pontine stroke using a 1,024-word vocabulary with a 25.5% word error rate (WER) at 78 words per minute (WPM) ^10^.

Another promising approach to restoring communication to people with dysarthria is through intracortical BCIs (iBCIs), which use penetrating microelectrodes to harness signals at the level of individual neurons from small regions of cortex. While iBCIs have achieved highly accurate brain-to-text decoding in people with amyotrophic lateral sclerosis (ALS), down to a 0.8% WER at 30.6 WPM with a 125,000-word vocabulary ^11–15^, it remains unclear whether their performance benefits will translate to dysarthria secondary to brainstem stroke.

In people with pontine stroke, MRI studies have shown reduced intra-network functional connectivity and decreased cerebral blood flow in precentral gyrus (PCG) ^16–18^. Structural imaging further demonstrates cortical thinning in these regions ^19–21^. Cortical thinning and disruptions in functional connectivity may alter the depth or spread of word-related information in speech-related areas in PCG, questioning whether intracortical microelectrode arrays can reliably target and record these signals from small regions of cortex in people with chronic pontine stroke to enable effective brain-to-text communication.

In this work, we demonstrate the potential utility of brain-to-text iBCIs for people with long-standing dysarthria and tetraplegia secondary to pontine stroke using as few as one 3.2 x 3.2 mm intracortical microelectrode array targeting speech-related precentral gyrus. Using both standard performance assessments and conversational tasks, we show that iBCIs can achieve substantially improved brain-to-text error rates compared to previous BCIs in our participants with a chronic pontine stroke. We then blueprint the data requirements to achieve high-performance brain-to-text in our participants, and we show that we can adjust for the effects of signal nonstationarities in new sessions by fine-tuning our brain-to-text decoder with as few as 22 new training sentences for T16 and 41 new training sentences for T22. Finally, we characterize how nonstationarities affect word representations across sessions and show that while representations remain informative, there are substantial across-day instabilities. Altogether, this work expands the applicability of state-of-the-art brain-to-text iBCIs to include people with brainstem stroke.

## Materials and Methods

### Clinical trial participants

This study includes data from participants T16 and T22 enrolled in the BrainGate2 clinical trial (ClinicalTrials.gov identifier NCT00912041). The trial was approved under an Investigational Device Exemption (IDE) by the US Food and Drug Administration (#G090003; CAUTION: Investigational device. Limited by federal law to investigational use.) and by the Institutional Review Boards at Mass General Brigham (protocol #2009P000505), Brown University (protocol #0809992560), the VA Providence Healthcare System (IRB-2011-009), and Emory University (protocol #00003070). T16 and T22 gave informed consent prior to any experimental procedures. All research was performed in accordance with relevant guidelines and regulations.

T16 is a right-handed woman with tetraplegia and dysarthria secondary to a pontine stroke approximately 19 years prior to enrollment in the clinical trial. We placed four 64-channel silicon intracortical microelectrode arrays (“NeuroPort” electrode arrays; 1.5 mm electrode length; Blackrock Neurotech, Salt Lake City, Utah, USA) in T16’s left precentral gyrus, guided by individualized Human Connectome Project (HCP) multimodal cortical parcellation (see *Array placement targeting* section below) ^22^: one in HCP-identified ventral premotor cortex (6v), one on the border of the HCP-identified premotor eye fields (PEF) and speech-related area 55b, and two in HCP-identified hand knob area (6d). T16 is able to speak slowly and quietly, but her speech production is severely impaired by limited facial, mouth, and diaphragm control. In our experience, her speech is unintelligible to new listeners, requiring more than a week of frequent conversation to understand 50% of her speech without repetition. She has limited voluntary control of her upper extremities, with some shoulder motion and some slow and contractured wrist and finger movements. She has limited to no voluntary control of her lower extremities. T16’s sensation is fully intact. Data for this study was collected with T16 between post-implant days 32-736.

Participant T22 is a woman with a history of brainstem stroke that occurred approximately three years prior to enrollment. She has anarthria and tetraparesis at baseline, and communicates through a combination of silent orofacial gestures and an eye-gaze tracking system. Following a task-based and resting-state fMRI localization procedure to identify relevant anatomical targets for sensor placement, she underwent implantation of six 64-channel Neuroport microelectrode arrays in the left hemisphere. Five arrays were placed in the precentral gyrus (with one targeting Brodmann’s area 6d, three targeting area 55b, and one targeting area 6v) and one array was placed in the inferior frontal gyrus (IFG).

### Array placement targeting

Prior to microelectrode array placement, T16 and T22 completed MRI sessions to guide surgical array placement. We used a different targeting strategy for each participant. For T16, we targeted arrays using an individualized Human Connectome Project (HCP) multimodal cortical parcellation ^22^ derived from structural and resting-state functional MRI (Fig. 1A–F). For T22, we targeted arrays using task-based fMRI activation maps, without HCP parcellation (Fig. 1H–J). Additional array placement targeting details can be found in the supplementary methods.

**Fig. 1.**
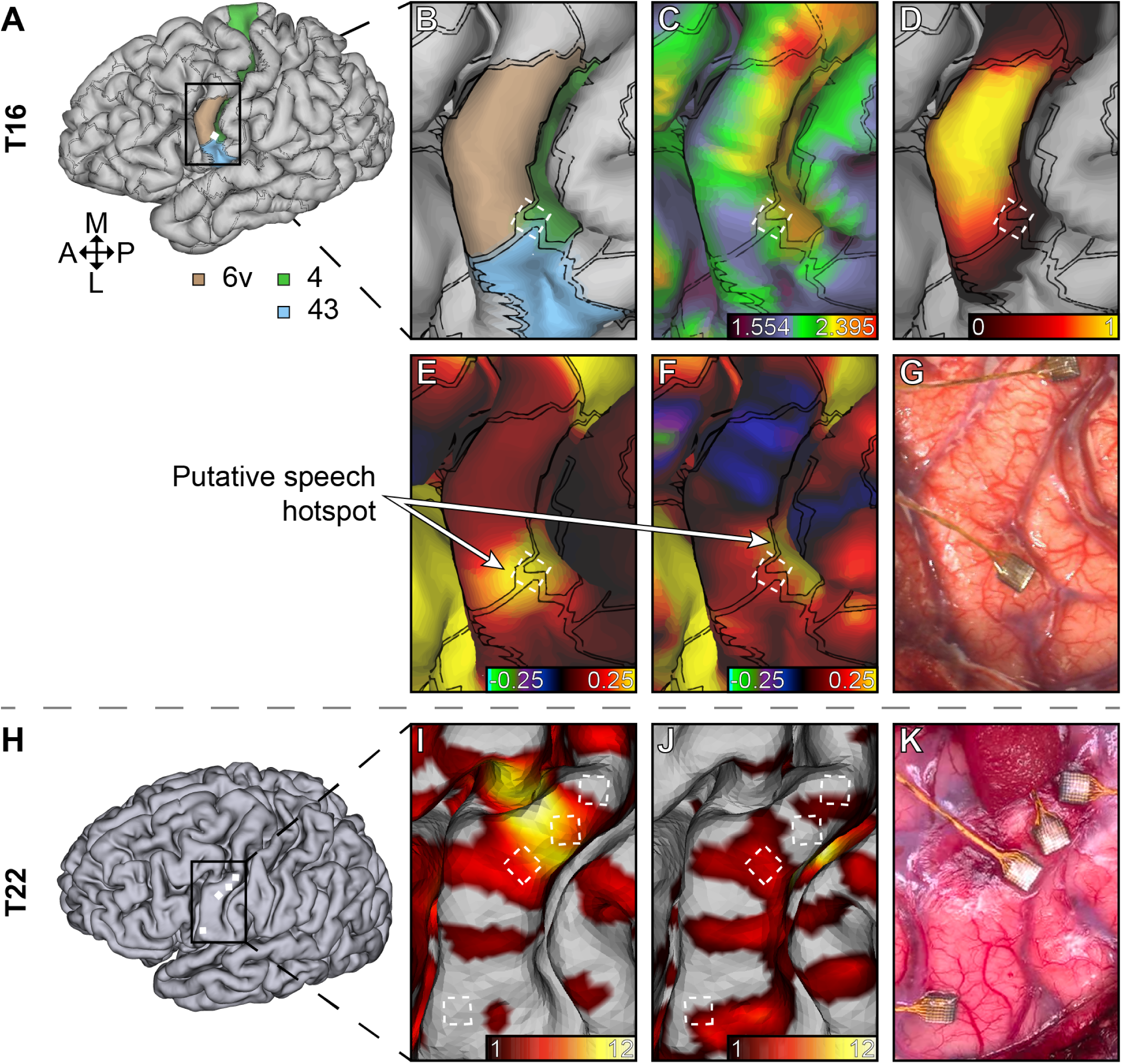
T16 (HCP parcellation-based) and T22 (task fMRI-based) pre-operative targeting and post-operative localization. **A**-**F** represent pre-operative images used to plan T16’s array placement with array border rectangles overlaid. (**A**) Pial surface of T16’s brain parcellated using the Human Connectome Project (HCP) multimodal cortical parcellation pipeline ^22^. Anatomical directions are consistent through **A**-**K**. (**B**) Zoomed-in region of the parcellated brain surrounding the 6v array target. (**C**) Same as **B**, but with the myelin map (T1w/T2w intensity ratio) projected onto the pial surface. Note the relatively lower myelin values in area 6v versus adjacent area 4. (**D**) Same as **C**, but a probabilistic heatmap representing the probability of a given surface area belonging to area 6v. (**E**) Same as **D**, but the group-averaged language-related resting state network number 25. Yellow and green indicate high and low resting state network connectivity, respectively. (**F**) Same as **E**, but the individual language-related resting state network number 25. (**G**) Intra-operative photograph of T16’s 6v array placement. **H**-**J** represent pre-operative images used to plan T22’s array placement with array border rectangles overlaid. (**H**) Pial surface of T22’s brain. (**I**) Zoomed-in region of the activation map from the sentence completion task. The color bar represents T-values. (**J**) Same as **I**, but for the orofacial motor (lips pucker/unpucker) task. (**K**) Intra-operative photograph of T22’s array placements.

### Data collection

Research sessions were performed at each participant’s residence. During each T16 session, the participant sat in a reclined chair in front of a computer monitor and was asked to perform different speech-related tasks in “blocks” lasting between 5 and 45 minutes each. Supplementary Table 2 lists all research sessions and number of blocks, as well as the attempted speech imagery T16 used during each session. In early sessions, we found that T16 quickly fatigued when vocalizing her attempted speech, so she switched to a more comfortable mimed strategy in which she mouthed words without producing sound, as was done in previous brain-to-text iBCI studies ^11–13,15^. After the 31st session, T16 voluntarily slowed her miming speed to subjectively improve word accuracy. For each T22 session, the participant laid in her bed with her head and trunk slightly elevated. Tasks were displayed on her television wall-mounted at the foot of her bed. Supplementary Table 3 lists all research sessions during which we collected data with T22. Systems used for data collection are described in the supplementary methods.

### Neural signal processing

We recorded from the electrode arrays with the NeuroPort System (Blackrock Neurotech) and processed signals as in prior brain-to-text iBCI studies ^11,12^: analog band-pass filtering, digitization at 30 kHz, linear regression referencing (LRR) ^23,24^, and digital high-pass filtering at 250 Hz. On each channel, we computed threshold crossings (−4.5 times root-mean-square voltage for T16; −3.5 times for T22) and spike-band power (SBP) in 20 ms bins.

For T16, only the 64 channels from the 6v array were used for brain-to-text decoding. We excluded features from the 55b array due to little phonemic tuning and substantial modulation to other behaviors ^25^. For T22, the 64 channels from the 6v array, and the 64 channels from each of three 55b arrays (for a total of 256 channels) were used in this study. T22’s IFG and 6d arrays were excluded due to little phonemic tuning.

### Task descriptions

#### Copy task

Each trial began with a delay period with an on-screen written cue, with participants instructed to read the cue and prepare to attempt to say it. To compare performance across different vocabularies, participants were cued with sentences from either a 50-word vocabulary ^9^ or from the Switchboard corpus representing a large vocabulary ^26^.

#### Diagnostic task

During the delay period, the task cued one of eight speech conditions: “bah”, “choice”, “day”, “kite”, “though”, “veto”, “were”, and “---” (an instruction to do nothing) ^11,12^. After 0.5 s, the cue descended at a constant speed and flashed green when centered in a gray rectangle approximately 2 s later (execution), cueing T16 to attempt to say the word or do nothing. During inter-trial periods, participants were instructed to rest. Illustrations of the copy and diagnostic tasks are presented in Fig. 3A and 5A, respectively.

#### Question-and-answer (Q&A) task

Q&A task blocks prompted T16 to sequentially answer one or more questions using the brain-to-text iBCI. Each block began by displaying a question either about T16’s interests or a general conversation starter. When T16 finished planning her response, she indicated to the system operator with a nod to initiate the response period, then attempted to speak as we displayed the iBCI’s predicted text. Upon sentence completion, she nodded to the system operator, initiating a verification stage in which T16 vocalized the “ground truth” sentence she attempted to say as the operator recorded it. Then, she indicated to the operator when she was ready to attempt her next sentence, repeating the response period until she completed her response to the question.

After Q&A sessions, we reviewed video footage to verify or correct the transcribed ground truth sentences via lip reading, and excluded any sentences in which we were not 100% confident in our post-hoc transcription accuracy. We also excluded any sentences during which T16 intentionally repeated a missed word, as a language model would skip grammatically incorrect word repetitions, or T16 paused to reread previously predicted text, due to evidence of reading-related neural modulation ^13^. After offline verification, 19 sentences were excluded, leaving 102 verified sentences analyzed in response to 29 questions.

For all tasks, we excluded trials with loud ambient sounds during the execution period to avoid listening-related neural modulation ^13^ and any trials interrupted by baseline, intermittent behavioral anomalies, such as stretching or muscle spasms, if they occurred during the execution period. Additional task details are in the supplementary methods.

### Brain-to-text decoding

Brain-to-text decoding, illustrated in Fig. 2A-D, followed previously published iBCI studies ^11–15^. Non-speech electrodes were zeroed (T16) or dropped (T22), resulting in a 512-dimensional neural feature vector of 20-ms binned threshold crossings and SBP for each participant that was z-score normalized and smoothed using a 40-ms Gaussian kernel. The first stage of the model was a recurrent neural network (RNN) phoneme decoder that was trained using the connectionist temporal classification (CTC) loss to predict phoneme probabilities from neural features. The RNN consisted of session-specific 512-unit input layers which accounted for neural drift across sessions that fed into five gated recurrent unit layers with 512 tanh units each. A dense linear output layer then produced phoneme probabilities for 41 phoneme classes every 80ms.

**Fig. 2.**
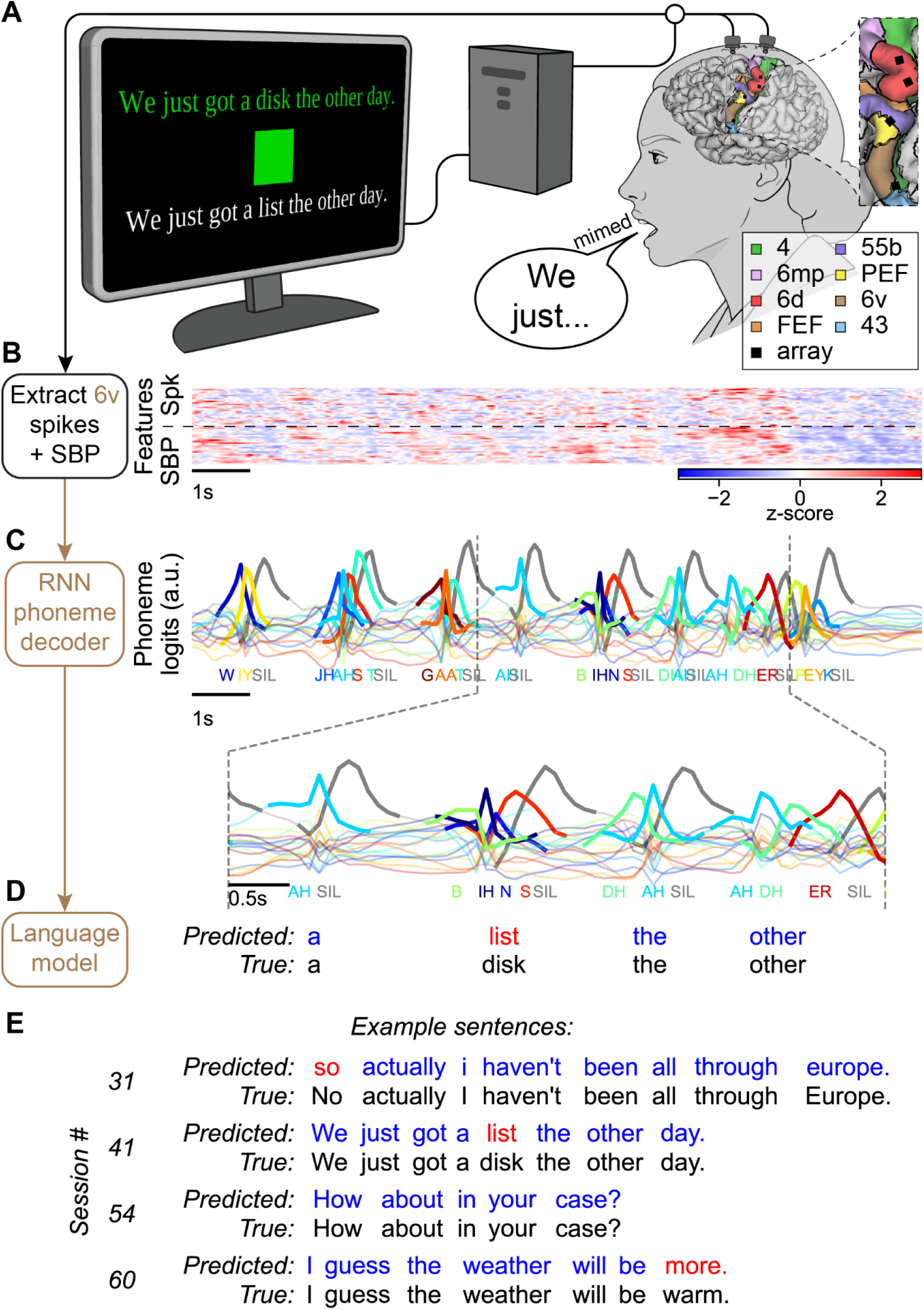
Brain-to-text decoding in a person with pontine stroke. (**A**) T16’s cortical surface reconstruction with array placements and task description. During the copy task, T16 was prompted with a sentence to mime (mouth words without vocalization). The brain-to-text intracortical brain-computer interface (iBCI) predicted each sentence as T16 mimed. Data in **B**-**D** represent T16 miming the sentence, “We just got a disk the other day.” (**B**) Example normalized threshold crossing rate (Spk) and spike band power (SBP) features extracted from T16’s 6v array while miming during the copy task on post-implant day 428. (**C**) A trained recurrent neural network (RNN) predicted phoneme probabilities over time as T16 mimed each sentence. Labels representing phoneme sounds are displayed below the time points at which they were predicted. For visual clarity, only the 17 predicted phonemes’ probability sequences are plotted here. Inset shows a zoomed portion of the predicted phoneme probability sequence. (**D**) A series of language models converted phoneme probability sequences representing each mimed sentence into a corresponding sentence prediction (see *Methods*). (**E**) Four example predicted and ground truth sentences are shown. Four sessions were selected that most closely match the median 125,000-word vocabulary word error rate (WER) across all sessions (Fig. 3C). One sentence was selected per session that best matched the median WER for that session. Blue or red words were correctly or incorrectly predicted, respectively.

**Fig. 3.**
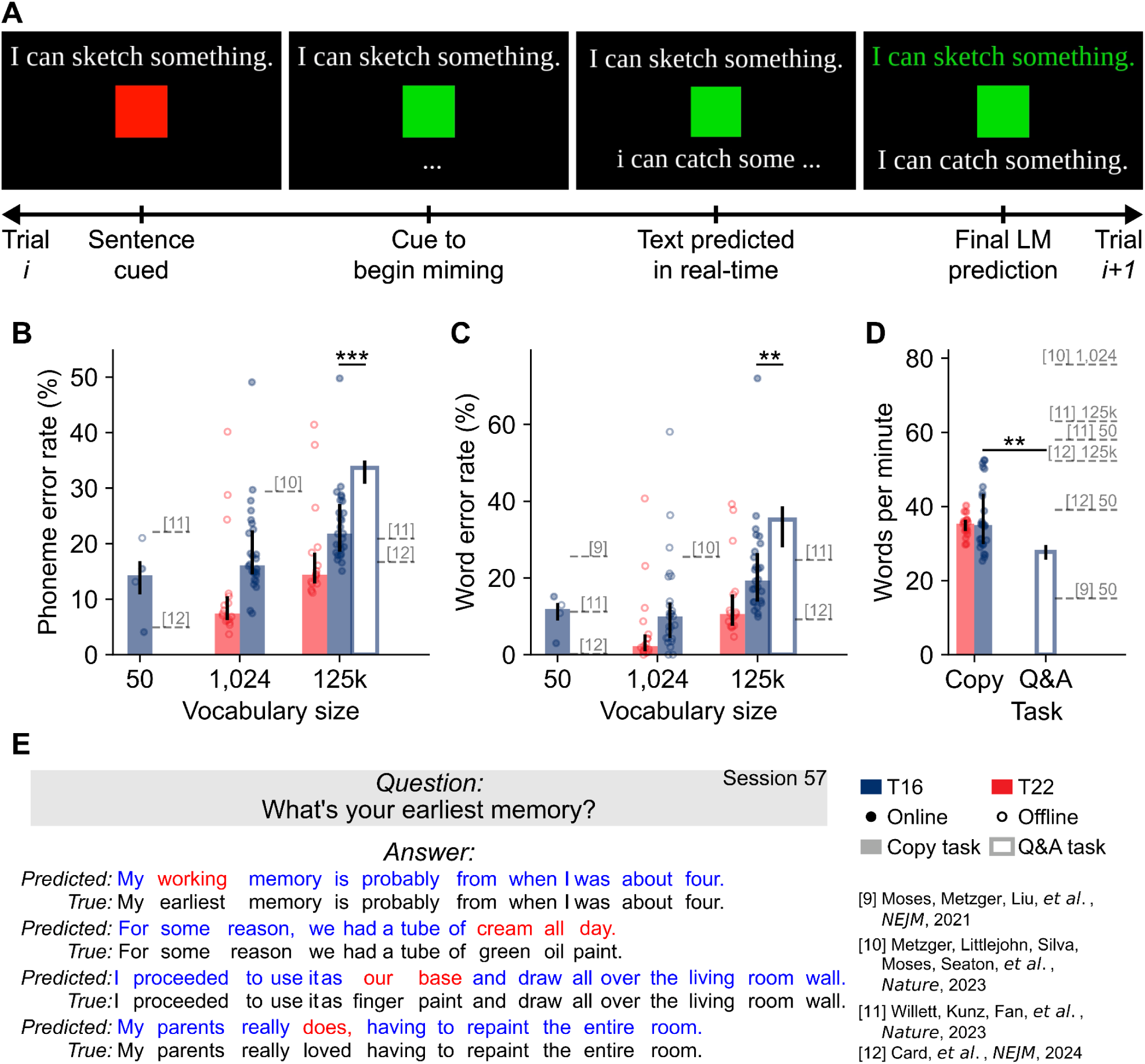
Highly accurate brain-to-text decoding with intracortical microelectrodes in pontine stroke. (**A**) Illustration of the copy task. A sentence was prompted over a red square, instructing participants to read the sentence but not mime it. Then, the square turned green, cueing them to begin miming the sentence. For T16, as she mimed the sentence, the brain-to-text iBCI predicted what she mimed. After T16 completed miming the sentence, a researcher pressed a button to instruct the language model to predict the final sentence given the full phoneme probability sequence. For T22, there was no closed-loop evaluation, so no on-screen predictions were displayed. (**B**) Session phoneme error rates for sentences composed of varying vocabularies. Blue represents T16, and red represents T22. Markers represent individual sessions, bars represent median across sessions, and thin lines around the median bar represent 25th to 75th percentiles across sessions. Filled markers represent sessions with held-out closed-loop evaluation blocks, and empty markers represent sessions with offline evaluation trials (that did not have closed-loop evaluation blocks). Filled bars represent the copy task, and empty bars represent the question and answer (Q&A) task. Dashed lines represent mimed (also referred to as “silent” in source text) metrics reported in previous intracortical or electrocorticography (ECoG) studies, if reported. If mimed metrics were not reported, vocalized metrics are presented. For all comparisons in **B**-**D**, ** represents *p* < 0.01, and *** represents *p* < 0.001 by a two-sided Wilcoxon rank-sum test. (**C**) Same as **B**, but WERs. (**D**) Same as **C**, but words predicted by the iBCI per minute. Copy task markers represent 125,000-word language model closed-loop evaluation blocks. Dashed lines citing prior studies include the language model vocabulary sizes used for decoding. (**E**) An example of the Q&A task with T16 with exemplary responses to the question, “What’s your earliest memory?”

After phoneme decoding, 5-gram language models (LMs) were used to select the most likely word sequence from the predicted phoneme probabilities. The 50-word and 125,000-word LMs used previously published parameters ^11,27^; we trained the 1,024-word LM with the same methods ^11^ on OpenWebText2 ^28^ sentences that contained only words from the 1,024-word vocabulary used in ECoG studies ^10^. For the 125,000-word and 1,024-word LMs, a final prediction at the end of each sentence rescored the 100 best 5-gram candidates with a 6.7B-parameter open pretrained transformer LM ^29^, as done previously ^11,12^.

We primarily used three metrics to assess brain-to-text decoding performance: words per minute (WPM), phoneme error rate (PER), and word error rate (WER). We computed WPM as the total number of words predicted by the iBCI divided by the duration of the execution periods. We computed phoneme error rate and word error rate as the Levenshtein distance summed across all evaluation sentences of a task within a session, divided by the total number of ground-truth phonemes or words consistent with prior work ^11,12^.

Complete training details are provided in supplementary methods and all hyperparameters are listed in Supplementary Table 5.

### Diagnostic task analysis

To investigate word-level representations of intracortical recordings, we analyzed diagnostic task data (seven words and a do-nothing condition, 10 trials each per session). We smoothed binned spike counts using a 120-ms Gaussian kernel and we aligned neural activity to -3.5 s to 2.0 s relative to the execution period when T16 mimed the cued word (if not instructed to do nothing). Peri-stimulus time histograms (PSTHs) showed the average and standard error of the mean response to each condition.

### Linear discriminant analysis training, adaptation and evaluation

We assessed the stability of population-level speech representations in T16 using the diagnostic task, which we collected approximately weekly, allowing us to evaluate how the observed neural representations of specific words evolved over time.

We trained a linear discriminant analysis (LDA) classifier to predict which word T16 mimed during each trial. For every session, the smoothed binned spike counts and SBP from each 6v array electrode were first averaged per feature in a window from 0.5 s before to 1.0 s after the execution cue, a window selected empirically based on PSTHs (see Fig. 5B, C) to capture miming-related modulation. We then reduced the time-averaged neural features from each trial using principal component analysis (PCA) to prevent overfitting, then fit an LDA to maximize between-class separability relative to within-class variance. The number of principal components retained for LDA was chosen through five-fold cross-validation to maximize classification accuracy and minimize overfitting. We found that 25 components reliably yielded peak classification accuracy and this was used to train and evaluate all classifiers.

To assess cross-session generalization, each single-session’s PCA and LDA mappings were directly applied to data from all other sessions without retraining. The PCA and LDA weights and biases for each reference session were computed via averaging the weights and biases from each cross-validation fold.

LDA classification accuracy was computed as the total number of trials with a correctly predicted class label divided by the total number of trials, and chance performance was computed by shuffling the channels input into the LDA classifier for a given session.

### Data recommendations sweep

To estimate data required for model training to achieve certain brain-to-text performance, we conducted three sweeps, varying: 1) the number of training sentences from a single session (Fig. 4A, Supplementary Fig. 1A), 2) the number of sessions and the number of finetuning sentences from the final evaluation session (Fig. 4B, D, Supplementary Fig. 1B, D), and 3) the time between decoder training and evaluation without recalibration (Fig. 4C, Supplementary Fig. 1C). All available sentences were used for training and validation, but only Switchboard and 50-word vocabulary sentences were used for evaluation. We fit exponential curves to all data points across all evaluation sessions to estimate the relationship between data quantity and PER. We inverted these exponential curves to estimate the data quantity recommended to achieve a selected PER, with standard deviations computed via the delta method ^30^. See supplementary methods for additional details.

**Fig. 4.**
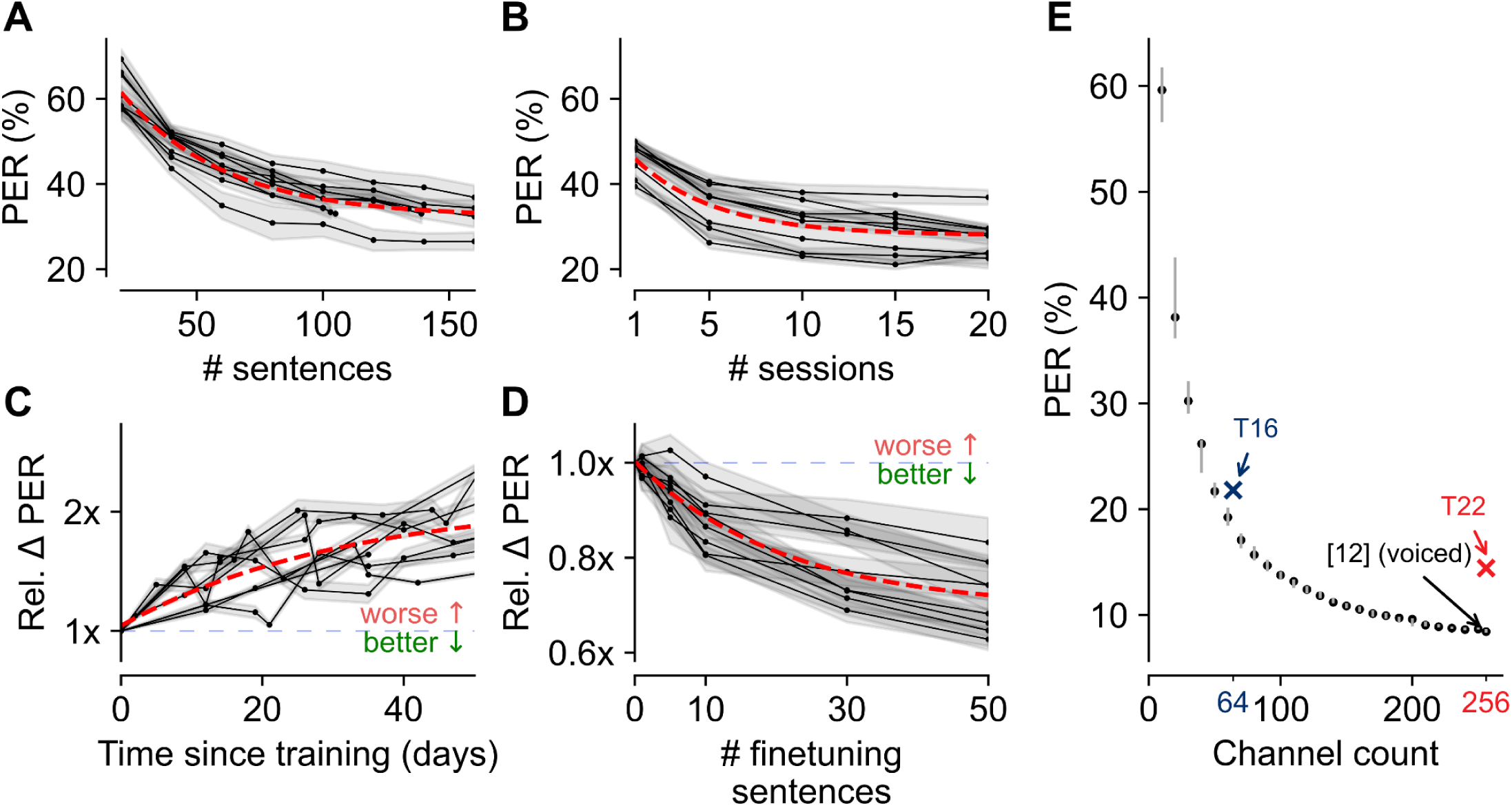
Scaling properties for brain-to-text decoding from intracortical microelectrodes in pontine stroke. (**A**) PERs resulting from training a phoneme decoder on a select number of calibration sentences from a single T16 session. Eight sessions are shown. The red dashed trace represents an exponential fit to all data points. Dots represent evaluation points. (**B**) PERs resulting from varying the number of calibration sessions with 50 calibration sentences from each T16 session. (**C**) Relative change in PER from frozen models evaluated on sentences from future T16 sessions. Relative change in PER is computed as the ratio between the PER on sentences from a future session and the PER on heldout calibration sentences from the last calibration session. (**D**) Relative change in PER resulting from finetuning a previously trained model on a small set of calibration sentences from a new T16 session. Relative change in PER is computed as the ratio between the PER after finetuning on the indicated number of sentences relative to the PER without any finetuning. In A–D, traces represent the mean PER across five random seeds and shaded regions represent the standard error of the mean across seeds. (**E**) Tradeoff between PER and channel count, copied with permission from ^12^. The blue X indicates T16 median performance from Fig. 3B with 64 channels. The red X indicates T22 median performance from Fig. 3B with 256 channels. Dots with whiskers represent median, 25th, and 75th percentiles of performance across random samples of channels.

## Results

### High accuracy brain-to-text from intracortical recordings with pontine stroke

To investigate whether iBCIs can restore naturalistic speech-based communication in a person with pontine stroke, we worked with participants T16 and T22, who have long-standing tetraplegia and dysarthria secondary to a pontine stroke. T16 speaks slowly and quietly with limited orofacial control, and T22 has orofacial movement but cannot phonate; therefore, for this study, both participants mimed all sentences without laryngeal vocalization except where noted. We found that 56 ± 3.1 of T16’s 64 channels in area 6v and 242 ± 8.8 of T22’s 256 channels in areas 6v and 55b showed substantial modulation during attempted speech (example for T16 in Fig. 2B).

To enable brain-to-text communication, we trained an RNN decoder to predict phoneme probabilities (see *Methods*) as participants mimed sentences from the Switchboard corpus used in prior brain-to-text iBCI work (Fig. 2C) ^11–15,26^. Then, we used a series of language models to convert phoneme probability sequences into the highest likelihood word sequence from a 125,000-word vocabulary (Fig. 2D). Example predicted sentences from four T16 sessions are portrayed in Fig. 2E.

We used a copy task (illustrated in Fig. 3A) to prompt participants with sentences from the Switchboard corpus, enabling rigorous performance assessment and comparison to past studies. All decoders were evaluated on separate blocks in which the model was held frozen with no further training (Fig. 3B-D, filled circles).

Across all 125,000-word vocabulary evaluation sessions for this work, which spanned 3 months to 2 years post-implant with T16 and 71 days for T22, T16 mimed sentences at a median 35.0 words per minute (WPM) and T22 at 35.3 WPM (Fig. 3D). Evaluating phoneme prediction (before any language models), phoneme error rates (PERs) were median 21.8% across sessions for T16 (closed-loop) and 14.4% for T22 (offline; Fig. 3B). This performance corresponded to a median word error rate (WER) of 19.4% for T16 and 10.6% for T22 at the final output after all language models (Fig. 3C; see Supplementary Table 6 for a summary of all performance metrics and comparisons to prior work).

To compare to prior literature, we also evaluated brain-to-text performance on 50-word *(9*; T16 only) and 1,024-word ^10^ vocabularies. With a language model trained and evaluated on sentences from a 50-word vocabulary, we achieved a median PER of 14.3% (Fig. 3B), corresponding to an 11.9% WER (Fig. 3C), a 53.5% reduction in WER compared to prior ECoG work in pontine stroke ^9^. To compare to previously presented results for a 1,024-word vocabulary ^10^, we used the closed-loop phoneme probability sequences predicted by a decoder trained on a large vocabulary ^26^, but only for sentences containing words exclusively from the 1,024-word vocabulary. Then, we used a 1,024-word language model to predict the sentences offline. This approach represents a conservative comparison because the RNN was trained on sentences from the larger Switchboard corpus and could not optimize for the more limited vocabulary. This resulted in a median closed-loop PER of 16.1% for T16 and offline 7.43% for T22 (Fig. 3B) and a 10.0% offline WER for T16 and 2.25% for T22 (Fig. 3C), an average 76.0% reduction in WER compared to prior ECoG work in pontine stroke ^10^ with as few as one NeuroPort electrode array placed in a putative speech area.

### Restoring rapid conversational brain-to-text in pontine stroke

An important real-world application of brain-to-text iBCIs is the ability to engage in a conversation. To demonstrate the brain-to-text iBCI can translate to conversational usage with T16, we prompted her with free response questions. She mimed each sentence of her responses as the brain-to-text iBCI predicted the sentences in real time. After each sentence finished decoding, we asked T16 to vocalize the sentence she just said to serve as a ground truth reference. In this Q&A task conducted in 8 sessions, T16 used the brain-to-text iBCI to say 102 sentences in response to 29 questions at 27.7 WPM with a PER of 33.7% and a WER of 35.2% (Fig. 3B-D). An example set of response sentences is shown in Fig. 3E.

### Decoder fine-tuning enables rapid brain-to-text decoding

To gauge the up-front time investment for a putative new user to achieve usable brain-to-text iBCI, we trained models on varying numbers of Switchboard corpus calibration sentences from a single session (Fig. 4A and Supplementary Fig. 1A). We observed that PER improved as sentence count increased and appeared to follow an exponential relationship, consistent with prior work ^11,12^, so we fit an exponential curve to all pairs of sentence counts and PERs across eight sessions for T16 and seven sessions for T22 (see *Methods*). We used the exponential fit (see *Supplementary Methods*) to estimate the number of calibration sentences to reach 95% of the best theoretical single-session performance. On average, T16 required just 123 ± 13.1 sentences (21.4 ± 2.28 minutes at T16’s copy task miming rate) while T22 required only 116 ± 4.5 sentences (20.0 ± 0.78 minutes at T22’s copy task miming rate) to achieve 95% of the estimated best single-session PER (equivalent to 34.6% PER for T16 and 30.7% PER for T22). Furthermore, with data from multiple sessions, we found PER could be further reduced to 29.1% for T16 and 29.7% for T22 (95% of the estimated minimum for the exponential curve fit to PERs vs. session counts) by incorporating 13 ± 3.6 sessions for T16 and 7 ± 1.3 sessions for T22 with 50 sentences each (Fig. 4B, Supplementary Fig. 1B).

Prior work has shown that decoders trained on one day require recalibration to maintain or improve performance on future days ^31–35^. Without recalibration, PER of decoders trained on all sentences from all previous sessions on average had a 25% relative increase in phoneme error rate by 6.9 ± 1.8 days after training for T16 (Fig. 4C) and for T22, PER also rose and the increase scaled with the number of training sessions (Supplementary Fig. 1C). To compensate for this, we applied a finetuning approach used in prior brain-to-text iBCI work in clinical trial participants with ALS to restore performance using a small set of calibration sentences from the new session ^12^. With just 22.4 ± 2.7 new calibration sentences (3.90 ± 0.47 minutes) for T16 and 40.7 ± 2.8 new calibration sentences (7.03 ± 0.48 minutes) for T22, we could finetune the phoneme decoder to achieve a 20% reduction in PER relative to the performance without any finetuning (equivalent to 65.1% (T16) and 49.3% (T22) of the maximum possible PER reduction), as shown in Fig. 4D and Supplementary Fig. 1D. In summary, we generally saw improved performance when incorporating more training data.

Another key component of brain-to-text performance is channel count, where prior studies have shown that more intracortical recording channels yields higher performance with a log-linear relationship ^11,12^. We compared the performance achieved with T16’s 64 and T22’s 256 speech-related channels against performance-versus-channel count relationships derived from people with ALS ^12^. Performance was in-line with those prior expectations (Fig. 4E), suggesting that intracortical interfaces in people with pontine stroke would benefit from channel scaling similar to that seen in people with ALS.

### Neural recordings remain informative despite channel-level instability

While our evaluations demonstrated accurate brain-to-text decoding over long time periods (more than two years post-implant), achieving this high accuracy required finetuning phoneme decoders to neuronal activity in each individual session. Investigating the source of this finetuning requirement was difficult using copy or question-and-answer task data, as the lack of consistent structure across trials and days made it challenging to evaluate neural representations at either the single-channel or population levels. Thus, to investigate cross-session model drift directly, we asked T16 to perform an alternate task that cued her to mime a word with precise timing or do nothing during each trial (diagnostic task, see *Methods*; Fig. 5A). This task enabled systematic analysis of neural representations via repeated collection of a small set of speech-related conditions. T16 completed the diagnostic task in 38 sessions spanning 420 days during the study, enabling rigorous assessment of neural recording stability over a long time period.

**Fig. 5.**
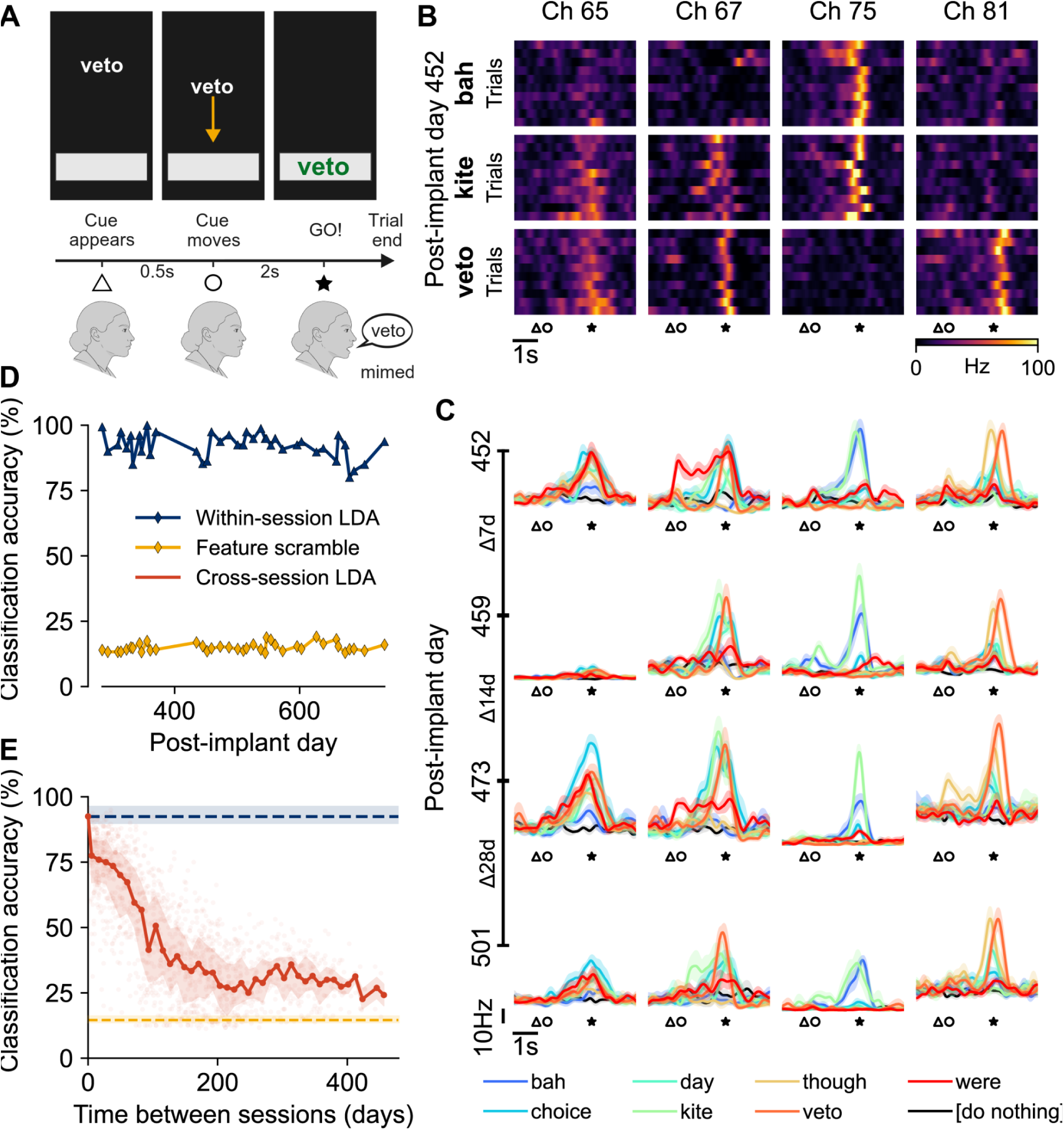
Neural feature stability over time with T16. (**A**) Diagram illustrating the diagnostic task (see *Methods*). Each trial began with one of eight conditions (seven words and “do nothing” represented by “---”) appearing at the top of the screen. After 0.5 s, the word began to scroll down the screen at a constant rate until it hit a gray box. Once centered in the gray box, the word flashed green, cueing T16 to mime the word with precise timing. (**B**) Single-trial smoothed threshold crossings (one row per trial) for example channels when T16 mimed “bah”, “kite”, or “veto”. (**C**) PSTHs for the same channels as **B**, but across four sessions separated by 1, 2, and 4 weeks, respectively. (**D**) Cross-validated linear discriminant analysis (LDA) classification accuracy for classifiers trained and evaluated on the same session. (**E**) Classification accuracy for decoders trained on each session and evaluated on each other session. Each semi-transparent red point represents classification accuracy for a single pair of train and evaluation sessions. The solid red trace and shading represent the median and 25th to 75th percentile performance, respectively, of all pairs within 5 days. The “Within-session LDA” trace and shading represent the median and 25th to 75th percentile performance, respectively, across all sessions from **D**. The “Feature scramble” trace and shading represent median and 25th to 75th percentile feature scramble performance, respectively, across all sessions from **D**.

Within a session, individual channels’ responses for specific words were highly consistent across trials (Fig. 5B), illustrating the within-session representational reliability that enabled phoneme decoder training and evaluation. Peri-stimulus time histograms (PSTHs), which demonstrate tuning and modulation depths to all words, showed some channels (e.g., channel #81) have fairly consistent word representations across sessions, and others vary substantially (e.g., channels #65 and #67) across sessions (Fig. 5C), consistent with prior work ^31,36–38^.

We next characterized consistency at the population level by using linear discriminant analysis (LDA) classifiers to predict each trial’s condition. Cross-validated within-session classification accuracies for all sessions were median 92.5% (Fig. 5D), substantially higher than the median 14.6% accuracy with the neural features scrambled, indicating that word representations remained highly informative throughout the study period. Next, we applied these single-session LDA classifiers to different sessions to assess across-time stability (Fig. 5E). We observed an immediate drop in classification accuracy to approximately 75.5%, but interestingly, this accuracy remained stable until a difference of approximately 40 days of separation. Differences greater than 40 days began to see a gradual decline in accuracy to 39.1% (200 or more days separated), which is still greater than scrambled-feature performance and suggests that some word information remains consistently represented over long time periods.

## Discussion

We showed that a brain-to-text intracortical BCI can restore conversational communication with a 125,000-word vocabulary to people with chronic dysarthria caused by pontine stroke. Relative to prior ECoG literature involving people with pontine stroke, recordings from small speech-related regions of precentral gyrus yielded improved word error rates in 1,024-word and 50-word vocabularies. We observed comparable performance to prior iBCIs in ALS using similar electrode counts when evaluated on a 125,000-word vocabulary. To combat neural instabilities, we found finetuning a previously-trained model on 22 calibration sentences for T16 and 41 calibration sentences for T22 from a new session to reduce PER by 20% relative to the non-finetuned decoder. Furthermore, we found decodability of a small set of words remained consistent over time, though representation of these words in the neural recordings changed over the course of the study.

Our study adds to a growing body of literature demonstrating that basic model adaptation techniques can maintain brain-to-text iBCI performance in new sessions despite neural instabilities ^12,15,39^, here achieving low PERs and WERs for over 736 days. However, the necessity of per-session recalibration implies that the features used for decoding drifted substantially over time. Similar to the conclusions of prior work in (upper extremity-related) motor cortex ^36,40^ and some prior brain-to-text iBCI work ^12^, our findings on within-session LDA classification accuracy showed that population-level structure supporting word discrimination in ventral (orofacial) motor cortex remains present despite the across-day performance instability of individual classifiers. These instabilities might stem from changes at the electrode-tissue interface, day-to-day changes in the participant’s interactions with the brain-to-text iBCI, genuine functional remapping of cortical articulatory representations, or underlying fluctuations in neuronal engagement during similar tasks that can’t be detected when sampling only a sparse sampling of the network engaged in the task. Understanding the primary sources of instability will be helpful towards developing adaptive decoders capable of maintaining alignment between neural representations and intended speech output over long-term brain-to-text iBCI use.

For people living with brainstem stroke, these results provide the first evidence, to our knowledge, that penetrating microelectrode arrays can extract sufficient neuronal information for a BCI to restore accurate and rapid speech-based communication. Despite cortical thinning in PCG associated with pontine infarcts ^19–21^, we found existing microelectrode technologies commonly used in iBCI studies (NeuroPort) captured high-quality neuronal signals from two people with pontine stroke for at least the duration of this study, over 736 and 71 days, respectively. Furthermore, word representations remained strong despite known reductions in functional connectivity secondary to pontine stroke ^16–18^. Altogether, since the known reduction in cortical thickness and functional connectivity did not impact brain-to-text iBCI decoding in T16 or T22, these results provide a proof of concept that intracortical BCIs may be applied more broadly to address communication and other challenges faced by people with pontine stroke ^3–6,41–44^.

Using the iBCI for conversational purposes is a promising demonstration that brain-to-text iBCIs may soon restore rapid and accurate communication broadly to people with dysarthria. Although PERs and WERs for the Q&A task were higher than the copy task, T16 was still able to use the system to freely respond to questions as she desired. We hypothesize that the reduction in performance may be due to a word distribution that differs from the Switchboard corpus used for model training and performance assessment, or that the neural representations of spontaneous speech differ from those during cue repetition. Regardless, we expect decoding performance during the Q&A task to improve by incorporating Q&A data in the training set ^15^ and by increasing electrode count beyond the nearly 60 active channels used here.

Importantly, in this work, we expanded the pool of people with dysarthria who may benefit from brain-to-text iBCIs to include individuals with pontine stroke. This raises a broader question: what other stroke-related deficits might be addressed with intracortical BCIs? In pontine stroke, cortical language networks may remain largely preserved, allowing communication to be restored by bypassing disrupted subcortical pathways. In contrast, cortical strokes that cause aphasia directly damage key language and speech-generating areas ^45,46^, limiting the applicability of current brain-to-text BCIs that depend on those preserved networks. However, advances in modeling distributed brain systems suggest a path forward ^47–50^. These models could eventually enable the development of prosthetic cortical networks that substitute lesioned brain areas and interface with remaining neural tissue through intracortical recording and stimulation ^51–53^. Such prosthetic networks could, in principle, restore functions beyond communication, including motor control after stroke-related paralysis. With continued progress in mapping and modeling the neural substrates of language, future BCIs could potentially reconstruct prosthetic language networks capable of restoring communication to anyone affected by severe speech impairment.

## Data Availability

All data required to reproduce these results will be publicly released upon acceptance in a peer-reviewed journal.

## Acknowledgments

The authors thank participants T16 and T22 and their families and care partners for their contributions to this research. The authors also thank Yvan Bamps, Haris Rashid, Colleen Spellen, Maryam Masood, and Dave Rosler for administrative and clinical research support.

This work was supported by NIH-NINDS/OD DP2NS127291, Simons Foundation as part of the Simons-Emory International Consortium on Motor Control (CP), NIH F32HD112173 (SRNT), NSF-GRFP 1937971, 2439564, NIH T32EB025816 (ALP), Burroughs Wellcome Fund Career Awards at the Scientific Interface #1361581 (NSC), NIH K23DC021297, AHA Second Century Early Faculty Independence Award 23SCEFIA1156586 (DBR), NIH U01DC019430 (JJ, DBR, LRH), Cerebral Palsy Alliance Research Foundation, and Department of Veterans Affairs Rehabilitation Research, Development, and Translation N2864C (LRH). The content is solely the responsibility of the authors and does not necessarily represent the official views of the National Institutes of Health, or the National Science Foundation, or the Department of Veterans Affairs, or the United States Government.

## Author contributions

SRNT, PID, BJ, and CP conceptualized the project. SRNT, PID, BJ, and JJJ designed the tasks and research sessions. SRNT, PID, BJ, BAR, and PHB conducted research sessions with participant T16. JJJ conducted research sessions with participant T22. SRNT, PID, BJ, QR, and BMK wrote data analysis code. SRNT, PID, and QR analyzed all session data and generated figures. SRNT, ALP, DRD, EYC, NAY, and CP conducted MRI analyses and targeted placement of T16’s arrays. ACP and DBR conducted MRI analyses and target placement of T22’s arrays. ALP and LNW performed post-op co-registration of CT and MRI scans to align T16’s array placement with resting state networks. NSC, SDS, and DMB wrote brain-to-text BCI code, advised brain-to-text decoding practices, and provided relevant data from ^12^. LRH is the sponsor-investigator of the BrainGate2 multi-site clinical trial. NAY and CP supervised research with participant T16. LRH and DBR supervised research with participant T22. SRNT, QR, and CP wrote the manuscript. All authors reviewed and contributed edits to the manuscript.

## Declaration of interests

CP has served as a consultant and research scientist for Meta (Reality Labs) and is a senior adviser for Inductive Technologies.

DMB is a surgical consultant for Paradromics Inc.

SDS is an inventor on intellectual property licensed by Stanford University to Blackrock Neurotech and Neuralink Corp. He has patent applications related to speech BCI owned by the Regents of the University of California, including IP that has been licensed to the neurotech industry. Stavisky is an advisor to Sonera and a consultant for Neuralink.

The MGH Translational Research Center has a clinical research support agreement (CRSA) with Ability Neuro, Axoft, Neuralink, Neurobionics, Paradromics, Precision Neuro, Synchron, and Reach Neuro, for which LRH provides consultative input. Mass General Brigham (MGB) convenes the Implantable Brain-Computer Interface Collaborative Community (iBCI-CC); charitable gift agreements to MGB, including those received to date from Axoft, Blackrock Neurotech, Neuralink, Paradromics, Precision Neuro, and Synchron, support the iBCI-CC, for which LRH provides effort.

The MGH Translational Research Center has a CRSA with Paradromics, for which DBR has provided consultative input.

EYC is a consultant for Neuralink Corp.

NSC is a visiting neuroengineer for Neuralink Corp.

All other authors declare no competing interests.

## Data availability

All data required to reproduce the findings in this study will be publicly available following publication. The dataset will contain intracortical neural signals recorded from participants along with detailed information about the behavioral tasks.

## Code availability

The code for reproducing the figures will be publicly available following publication.

## Declaration of generative AI and AI-assisted technologies in the writing process

We used generative AI (ChatGPT, Gemini, Claude) for four purposes:

1. To assist with code writing for speed and improved reliability
2. To generate the person representation in Fig. 2A
3. To assist with developing questions for the Q&A task
4. After complete initial drafting of the text, to suggest modifications to improve readability and language of the manuscript

After using these tools, the authors reviewed and edited all code and text as needed and take full responsibility for the content of the article.

## Supplemental Material

Supplementary Methods

Supplementary Tables 1–6

Supplementary Figure 1

References 54–65

## Supplement

## Supplementary methods

### Array placement targeting

First, we focus on targeting for T16’s 6v array. T16 underwent a multi-modal (structural and functional) MRI session approximately 45 minutes in duration to guide surgical array placement, based on the HCP cortical parcellation and previously demonstrated with other iBCI participants ^11,12,54^. Scans were acquired with a 3T Siemens Magnetom PrismaFIT scanner equipped with a 32-channel head coil. T1-weighted (T1w) and T2-weighted (T2w) scans were collected at 0.8 mm isotropic voxel resolution, while phase-reversed resting state fMRI (rsfMRI) scans and spin echo fieldmap scans had 2.0 mm isotropic voxel resolution. Full scan parameters are listed in Supplementary Table 1. MRI scans were processed and parcellations were produced using the QuNex Singularity container (version 0.98.0, except where noted below) ^55^. The QuNex study was set up with the mapping and parameter files, and a list of commands run can be found in our GitHub repository (to be uploaded post publication). Briefly, structural images were processed to perform AC-PC alignment, brain extraction, readout distortion correction, bias field correction, and MNI registration (hcp_pre_freesurfer). FreeSurfer’s recon-all then generated white matter and pial surface models (hcp_freesurfer), followed by alignment to the HCP atlas surface using cortical folding (MSMSulc) and myelin map generation (hcp_post_freesurfer). rsfMRI images were corrected for motion and readout distortions, and registered to the T1w image (hcp_fmri_volume). rsfMRI volumetric data were then mapped to the surface and smoothed using a geodesic Gaussian kernel (2mm FWHM) (hcp_fmri_surface). Spatial independent component analysis was run on the rsfMRI data to remove noise related to motion, physiology, and the MRI scanner: components were auto-classified into signal and noise (hcp_ica_fix) and manually corrected by an expert (author EYC; hcp_reapply_fix), before regression of the noise components from the data. The alignment of the participant’s surface models to the HCP atlas surface space was then refined beyond the initial cortical folding-based alignment using the additional resting state connectivity and myelin information (hcp_msmall, hcp_dedrift_and_resample which used QuNex version 0.97.3 due to a bug in version 0.98.0). The precise alignment of the multi-modal cortical surface registration was checked by comparing the participant’s rsfMRI network and myelin features to those of the group atlas for area 6v and its borders with areas 4, PEF, and 43.

For array targeting ^11^, the group parcellation map (210P) was overlaid on the native pial surface model (fs LR 32k space) in Connectome Workbench ^56^(versions 1.5.0 and 2.0.1; Fig. 1A, B) to obtain a participant-specific estimate of Brodmann’s area 6v (Fig. 1D). Group and individual maps of resting state network 25 (Fig. 1E, F, respectively), which includes language-related areas such as Broca’s area, were further used to localize the “speech hotspot” in ventral 6v identified previously ^11^.

For T16, the array location was estimated by manual landmark-based co-registration. The preoperative T1-weighted MRI was reformatted in Brainlab Elements (Brainlab AG, Munich, Germany) to identify the oblique slice that most closely matched the intraoperative photograph (Fig. 1G). Two fiducial points were placed at sulcal junctions visible in both the intraoperative image and the matched MRI slice, and the known physical dimensions of the microelectrode array (3.2mm x 3.2mm square footprint) were used as an internal scale. Distances from each fiducial to the four array corners were measured and transferred to MRI space, and the resulting points were minimally adjusted to respect the known array geometry (square layout and fixed dimensions). These finalized corner coordinates were then converted from 3D RAS to LPS space and imported into Connectome Workbench to visualize array location relative to group and individual resting state networks and probabilistic heatmaps of area 6v (Fig. 1B-F).

For T22, we used task-based fMRI to guide array placement. The T1-weighted (T1w) images were initially processed and the surfaces constructed using the FreeSurfer v8.1.0 ^57^. This processing included motion correction, skull stripping, segmentation of the subcortical structures, intensity normalization, surface tessellation, topology correction, inflation, registration, and parcellation. The fMRI data were then preprocessed using fMRIPrep ^58^. The BOLD time series were motion-corrected, slice-time corrected, and resampled onto their original, native anatomical space using FreeSurfer bbregister ^59^. The time series were registered to the standard MNI152NLin2009cAsym space and then resampled onto fsaverage ^57^, and grayordinates space ^60^. All resamplings were performed with a single interpolation step by composing all the pertinent transformations (i.e. head-motion transform, susceptibility distortion correction, and co-registrations to anatomical and output spaces). Volumetric resamplings were performed using ANTs and surface resamplings were performed using FreeSurfer mri_vol2surf. T22 performed sentence completion (Fig. 1I) and orofacial motor (lips pucker/unpucker; Fig. 1J) tasks during the scan, which we used to identify regions of PCG that activated during speech. The preprocessed task fMRI data were input to the FreeSurfer Functional Analysis Stream (FsFast) for surface-based smoothing with a 4 mm full-width-half-maximum kernel, and a generalized linear modeling (GLM) with respect to the task design. The resulting hot-spot maps represented the significance (-log(p-value)) for each task indicating the key region(s) that was involved in that task. We placed the 55b and 6v arrays according to these activation maps, avoiding vasculature. Fig. 1H-J show placement of T22’s arrays overlaid onto the task activation maps, and Fig. 1K shows T22’s intra-operative array placement.

### Data collection system

To collect data, we used dedicated computers running modular Python and C++ processes that executed tasks and synchronized with neural recordings using the Backend for Realtime Asynchronous Neural Decoding system (BRAND) ^61^. The mobile neuroprosthetic rig incorporated four networked computers. The first ran Windows 10 version 1809 to interact and control the neural signal processing system (see Neural signal processing methods below) and also served as the primary interface with which the iBCI operators interacted with the system. The second computer ran Ubuntu 22.04 LTS with Linux kernel 6.1.66 with real-time kernel patch version 19 and was responsible for all signal processing and task control for mimed attempted speech sessions or just signal processing for vocalized attempted speech sessions. A third computer ran Ubuntu 22.04 LTS and was responsible for all compute-heavy tasks, such as online model training and inference (see Online phoneme decoder finetuning and evaluation section below), and visualization. This third computer additionally managed task control for vocalized attempted speech sessions. The fourth computer ran Ubuntu 22.04 LTS and was responsible for the language models. See Supplementary Table 4 for other software version details. For T22, the tasks running on the second, third and fourth computers described above ran jointly on a single computer running Ubuntu 22.04.

During each T16 session, the participant sat in a reclined chair in front of a computer monitor (24.5” diagonal screen size, 1920 x 1080 pixel resolution, 280 Hz refresh rate) and performed speech-related tasks in a series of “blocks” lasting between 5 and 45 minutes each. For each T22 session, the participant lay in her bed with her head and trunk slightly elevated. Tasks were displayed on her television (55” diagonal screen size, 1920 x 1080 pixel resolution, 120 Hz refresh rate) wall-mounted at the foot of her bed.

### Additional task details

We used the copy task (Fig. 2A) to compare our brain-to-text performance to prior work by using the same corpora. To compare performance on the 50-word vocabulary, we used custom-written sentences containing words from a 50-word vocabulary ^9^. To compare performance on a large vocabulary, we used the Switchboard corpus that was sourced from conversational English ^26^. The set of Switchboard sentences used excludes those with violent or offensive topics, and further details regarding how the set of sentences was selected can be found in ^11^. Blocks of 4 to 53 trials used sentences all from the same corpus. Fig. 3A shows an example progression of the closed-loop version of the copy task. At the beginning of each trial, we prompted one sentence at a time at the top of the screen above a red square. This indicated the beginning of the delay period, during which time the participants were instructed to read but not yet attempt to say the sentence. After four seconds, the square turned green to indicate the execution period. During the execution period in online training and closed-loop evaluation blocks, we displayed the iBCI’s predicted sentence below the green square and updated it as the iBCI continued to predict new text from incoming neural activity. Open-loop training blocks did not have any text printed below the square. For the first three sessions, T16 pressed down on a 2 cm diameter button to indicate when she completed her attempt to say the sentence. Due to T16 experiencing difficulty pressing the button, in later sessions we changed to the system operator (a member of the research team) observing T16’s speech attempts and pressing the button when she completed each sentence. Upon pressing the button, the language model computed its final prediction (see Language model training and evaluation section below), which we then displayed on the screen and initiated synthesis of the text via a text-to-speech model ^62^. Once the audio finished playing, the task proceeded to a 2 s inter-trial period.

We used the diagnostic task to instruct T16 to repeat attempts to say a small set of words for many trials to enable analysis of neural modulation and drift. The eight speech conditions in the diagnostic task match those investigated previously: “bah”, “choice”, “day”, “kite”, “though”, “veto”, “were”, and “---” (an instruction to do nothing) ^11,12^. Throughout the diagnostic task, a 750 pixel wide by 100 pixel high gray rectangle was displayed with its lower edge 100 pixels from the bottom of the screen. At the beginning of each trial, one word was selected pseudorandomly (to ensure equal trial counts for all conditions) and displayed 482 pixels from the bottom of the screen, indicating the beginning of the delay period. After 0.5 s, the word began to fall down the screen towards the gray rectangle at 166 pixels per second. Once the word was centered within the gray rectangle, it flashed green to indicate the execution period when T16 should begin to attempt to say the word. After a 0.5 s inter-trial period, the next trial began.

The question-and-answer (Q&A) task assessed brain-to-text performance in a conversational context. A pool of questions covering T16’s interests and general conversation starters was prepared with the help of ChatGPT (OpenAI, San Francisco, CA, USA). Each block began by displaying a randomly selected, unused question above a red square while T16 read it and planned her response. When ready, she nodded to the system operator, who pressed a button; four seconds later the square turned green and the iBCI began predicting her attempted speech, displaying the predicted sentence below the square and updating it as new text was decoded. When she completed a sentence, she nodded again, the square turned blue, and the language model’s final prediction was displayed. T16 then vocalized the ground truth sentence, which the operator typed into a text box below the prediction and T16 verified. The operator pressed the button to clear the ground truth and, once T16 was ready, to return to the execution period with all previous sentences displayed. Blocks ended when T16 indicated she had finished responding to the question.

### Brain-to-text decoding

Brain-to-text decoding followed similar methods to previously published iBCI studies ^11–15^. As illustrated in Fig. 2A-D, phoneme logits were predicted by a phoneme decoder, followed by one or more language models converting phoneme logit sequences into sentences.

#### Neural signal processing

We used the NeuroPort Biopotential Signal Processing System (Blackrock Neurotech) to record from the NeuroPort electrode arrays. Voltage signals were analog band-pass filtered (0.3 Hz to 7.5 kHz, 4th order Butterworth filter), then digitized at 30 kHz (250 nV resolution). Then, we applied a digital high-pass filter (250 Hz, acausal 4th order Butterworth filter with 4 ms delay) for all sessions except for the first six sessions included in the study, which used a digital band-pass filter (250 Hz to 5 kHz, acausal 4th order Butterworth filter). We used linear regression referencing (LRR) with coefficients computed on the filtered signals from the previous block, then applied those coefficients prior to digital filtering ^23,24^. We computed LRR coefficients for the first speech-related block in a session on a reference block collected at the beginning of the session. During the first six sessions included in the study, we used an untimed version of the diagnostic task (see Additional task details above) to compute LRR coefficients, as was done previously ^11,12^. During all other sessions, we computed LRR coefficients on a 60 s block during which T16 was instructed to remain idle. For T22, all LRR coefficients were calculated from a 45 second block during which T22 was instructed to remain idle.

After rereferencing and filtering, we computed timings of threshold crossings on each channel. Thresholds used for a given block were set to -4.5 times the root-mean-square voltage of each signal during the final 60 s of the previous block, or the reference block for the first speech-related block in the session. For T22, thresholds were set to -3.5 times the root-mean-square voltage from just the reference block for all blocks. Then, we summed the number of crossings on each channel in 20 ms bins. We also computed spike-band power (SBP) for each channel by squaring the rereferenced and filtered voltages, then averaging in the same 20 ms bins as for threshold crossings. Only the 64 channels from the 6v array were used for brain-to-text decoding analyses in this study for T16. We excluded features from the 55b array due to little phonemic tuning and substantial representation to other behaviors ^25^. For T22, the 64 channels from the 6v array, and the 64 channels from each of three 55b arrays (for a total of 256 channels) were used in this study. T22’s IFG array and 6d array were excluded due to little phonemic tuning.

#### Feature processing

The phoneme decoder received a 512-dimensional neural feature vector as input, combining 20-ms binned threshold crossings and SBP from just the electrodes putatively placed in speech areas (Brodmann’s area 6v, plus area 55b for T22). Any features representing other electrodes were zeroed (for T16) or dropped (for T22) from the neural feature vector. During offline processing, all non-zeroed features were z-score normalized by mean subtracting within each block, then dividing by the session-wide standard deviation (after mean subtraction). During online processing (T16 only), all non-zeroed features were z-score normalized using statistics from the preceding 10-20 trials’ execution periods. Before being processed by the model, all normalized features were smoothed using a 40-ms Gaussian kernel.

#### Phoneme decoder architecture and training

We used a recurrent neural network (RNN) to predict phoneme probabilities from neural features. Neural features were input to day-specific dense input layers with softsign activation functions used to account for neural drift across sessions, followed by five gated recurrent unit layers with 512 tanh units each, then a dense linear output layer producing logits for 41 classes. The 41 classes consisted of the 39 American English phonemes, one “silence” class, and one connectionist temporal classification (CTC) blank class. To train and evaluate the RNN on sentence data, we converted sentences into phonemes using the Carnegie Mellon University pronouncing dictionary (http://www.speech.cs.cmu.edu/cgi-bin/cmudict) and the G2P-En Python package ^63^. The RNN output phoneme logits every 80 ms using the most recent 280 ms (14 bins) of neural features. During training, input neural features were dynamically augmented during each training batch to improve decoder generalizability and stability by adding white noise and constant offsets ^39^. To further regularize the model, we also applied dropout and L2 weight regularization during training. All hyperparameters noted here and listed in Supplementary Table 5 were verified to be optimal for T16 and T22.

#### Offline phoneme decoder training and evaluation

The offline phoneme decoder training protocol used here is similar to what was used previously ^11,12^. All offline RNNs trained for up to 10,000 batches of 64 sentences each, with early stopping based on validation phoneme error rate (PER; 3,000 batches; more details below). We used an adaptive moment estimation (ADAM) optimizer with a 0.01 initial learning rate, *β*1=0.9, *β*2=0.999, and *ε*=0.1. We applied learning rate annealing with a 0.9 decay factor, applied whenever validation CTC loss did not improve in 300 batches. Validation PER was computed every 50 batches, and model checkpoints were saved whenever validation PER had achieved a new low.

We pretrained RNNs offline that were later used for two purposes: 1) closed-loop evaluation sessions within the first 15 brain-to-text sessions (after online training as presented in Fig. 3, see Online phoneme decoder finetuning and evaluation section below; T16 only), and 2) offline analysis (for 50-word vocabulary for T16 and all results for T22 in Fig. 3 and all models used in Fig. 4 and Supplementary Fig. 1). Pretrained RNNs were trained on all sentences from all previous sessions, unless session count was experimentally limited (see Data recommendations sweep section below). For RNNs used online in session after finetuning (see Online phoneme decoder finetuning and evaluation section below), we held out 10 sentences per session for the validation set, and the remaining sentences were used for training. For RNNs used in offline analyses (all models used in Fig. 4 and Supplementary Fig. 1), we held out 10 sentences per session for the validation set, 10 other sentences for the evaluation set, and all remaining sentences were used for training (unless sentence count was experimentally limited, see Data recommendations sweep section below). During pretraining, training and validation batches consisted of sentences drawn from all sessions with equal probability.

For pretrained RNNs used in offline analyses (50-word vocabulary for T16 and all results for T22 in Fig. 3 and all models used in Fig. 4 and Supplementary Fig. 1), models were finetuned with data from the evaluation session. Finetuning began by loading a pretrained RNN’s lowest validation PER checkpoint, then copying the input layer weights for the most recent session (relative to the evaluation session) to a new input layer to accommodate the new session. Then, RNN finetuning proceeded the same as pretraining, with all weights trainable using the same hyperparameters (see Supplementary Table 5), except for two key changes: each training batch was composed of 60% evaluation session training sentences and 40% training sentences from previous sessions, and validation loss and validation PER were computed only on the evaluation session’s validation set (10 sentences). After finetuning, phoneme decoder performance was assessed on the 10 evaluation sentences from the evaluation session for all offline analyses except for the offline T22 results in Fig. 3 (both 125k and 1024 vocabularies), which instead used 30 evaluation sentences from the evaluation session to get a more robust evaluation.

#### Online phoneme decoder finetuning and evaluation

The online phoneme decoder finetuning protocol used here is similar to what was used previously ^12^, and was only used with T16. Online finetuning rapidly enabled high-performance phoneme decoding in session within approximately 30 minutes while keeping engagement high due to the closed-loop nature of the task. At the beginning of finetuning, we loaded a previously-trained RNN and all sentences used to train that pretrained RNN. Then, we prompted T16 with new sentences via the copy task, and updated model weights as T16 completed more trials. We used ADAM optimization with the same parameters as offline phoneme decoder training, except with a static learning rate of 0.004. Each finetuning epoch lasted between 32 and 200 batches of 64 sentences each, which stopped early if the average total loss (CTC and L2 regularization) over the previous 10 batches was below 0.5. Finetuning batches were composed of 60% sentences from the current session and 40% training sentences from previous sessions. We typically included data from between 3 and 20 previous sessions, noted in Supplementary Table 2. After a finetuning epoch completed, the RNN updated its weights used for inference during the subsequent inter-trial period, and a new finetuning epoch began.

During closed-loop evaluation sessions within the first 15 T16 brain-to-text sessions, we first pretrained models offline during the session using all previous sessions and the open-loop sentences collected so far in the evaluation session (see Offline phoneme decoder training and evaluation section above). Then, we loaded the pretrained model and performed online finetuning for approximately 50 sentences (see Supplementary Table 2 for number of online finetuning trials used).

After the first 15 T16 brain-to-text sessions, we reduced in-session training time by transitioning to exclusively online finetuning (no offline model training in session). In this paradigm, we started each session with the most recent prior session’s online-finetuned model. When we loaded its parameters, we copied the most recent session’s input layer’s parameters to a new input layer, assigned to the current evaluation session. Then, we proceeded with online finetuning as described above until WER had anecdotally plateaued for approximately 20 sentences. Due to a minor bug in our online finetuning code, for all closed-loop evaluation sessions between brain-to-text sessions 39 and 68 (10 total closed-loop evaluation sessions), input layers representing all sessions after session 38 were initialized with session 38’s input layer parameters, even if session 38 was not the most recent session relative to the evaluation session (in other words, we may have slightly reduced initial decoding performance using an older starting input layer). Model finetuning, including input layers, otherwise proceeded as described above.

To assess performance of online-finetuned models, we froze model parameters and evaluated performance on new sentences (see Supplementary Table 2 for number of evaluation trials in each session).

#### Language model training and evaluation

We used 5-gram language models (LMs) to select most likely word sequences from phoneme probability sequences predicted by phoneme decoders in a similar fashion to prior work ^11,12^. The 50-word and 125,000-word 5-gram LMs used previously-published parameters ^11,27^. Briefly, the 50-word 5-gram LM was trained on 2,413 custom-written sentences containing only words from the 50-word vocabulary ^9^. The 125,000-word vocabulary stems from the CMU pronouncing dictionary (http://www.speech.cs.cmu.edu/cgi-bin/cmudict) and encompasses the majority of English words, and its 5-gram LM was trained on the OpenWebText2 corpus ^28^. The 1,024-word vocabulary was derived from the NLTK Twitter and Cornell film corpora ^64,65^, and was used previously in brain-to-text work with ECoG ^10^. We trained the 1,024-word 5-gram LM using the same methods published previously for 50- and 125,000-word 5-gram LMs ^11^, but using only OpenWebText2 sentences in which all words belonged to the 1,024-word vocabulary.

During online finetuning and closed-loop evaluation, we predicted the most likely word sequence in real time as T16 mimed sentences using a pruned version of the 125,000-word LM, accelerating inference for low-latency word updates. Once T16 finished miming the sentence, we initiated a final prediction given the entire phoneme sequence, as was done previously ^11,12^. The 1,024-word LM for T16 and T22 only used this final prediction process, since it was only evaluated offline using phoneme sequences predicted from sentences containing words from just the 1,024-word vocabulary (during closed-loop evaluation for T16, or predicted offline for T22). To yield the final predicted word sequence, the pruned LM first predicted up to 100 of the most likely word sequences along with probability scores representing the likelihood of each sequence. Then, an unpruned version of the LM rescored the 100 candidate word sequences. Finally, the 100 candidate word sequences were separately scored by a 6.7B-parameter open pretrained transformer LM ^29^ that yielded its own scores for each word sequence. The transformer LM’s scores and the unpruned 5-gram LM’s scores were weighted and combined, and the final predicted word sequence was selected as that with the highest combined score. See Supplementary Table 5 for all LM hyperparameters.

We computed phoneme and word error rates using the Levenshtein distance, which counts the number of insertions, deletions, or substitutions to the predicted phoneme or word sequence to match the ground truth phoneme or word sequence, divided by the number of ground truth phonemes or words. Consistent with prior work ^11,12^, we aggregated Levenshtein distances and ground truth phoneme and word counts across all evaluation sentences for a given task type within a session to balance reported metrics across sentence lengths.

### Diagnostic task analysis

To investigate word-level representations of intracortical recordings, we analyzed data from the diagnostic task, in which T16 repeated a small, fixed set of seven words and a do-nothing condition, each repeated 10 times per session. We smoothed binned threshold-crossing counts using a 120 ms standard deviation Gaussian kernel and aligned neural activity from -3.5 s to 2.0 s relative to the execution cue. For four example channels, we plotted the aligned, smoothed activity of each trial of a given word (one row per trial) and peri-stimulus time histograms (PSTHs) showing the mean and standard error of the mean response to each condition on the same time axis.

#### Linear discriminant analysis training, adaptation and evaluation

We trained a linear discriminant analysis (LDA) classifier to predict which word T16 mimed during each trial. For every session, the smoothed binned threshold-crossing counts and SBP from each electrode within the 6v array were first averaged per feature in a window around the execution cue (0.5 s preceding to 1.0 s following) for each trial. This window was selected empirically based on PSTHs (see Fig. 5B, C) to capture visible miming-related modulation. We then reduced the collection of time-averaged, per-feature neural activity from each trial using principal component analysis (PCA) to prevent overfitting, followed by an LDA fit on the reduced data to maximize between-class separability relative to within-class variance. The number of principal components retained for LDA was chosen through five-fold cross-validation to maximize classification accuracy and minimize overfitting. We found that 25 components reliably yielded peak classification accuracy when averaged across sessions, so we used 25 components to train and evaluate all classifiers.

To assess cross-session generalization, each LDA classifier trained on one reference session was directly applied to data from all other sessions without retraining. The PCA and LDA weights and biases for each reference session were computed by averaging the weights and biases from each cross-validation fold, and all cross-session evaluations projected the new data through these fixed reference-session PCA and LDA mappings. LDA classification accuracy was computed as the total number of trials with a correctly predicted class label divided by the total number of trials, and chance performance was computed by shuffling the channels input into the LDA classifier for a given session.

### Data recommendations sweep

To estimate data quantity to achieve certain brain-to-text performance, we conducted three sweeps across various parameters controlling the amount of data available for model training. All T16 sweeps used eight slow mime speed sessions that were selected based on their ability to meet the trial count requirements of each sweep. All T22 sweeps used seven sessions with sentences from the Switchboard corpus. All Switchboard, OpenSubtitles (only T22), and 50-vocabulary (only T16) sentences collected in each session were available for training and validation, but only Switchboard and 50-vocabulary sentences were used for evaluation. For T16, all sentences were used regardless of whether they were collected as open-loop, online training, or closed-loop evaluation sentences in session. For T22, only open-loop sentences were used. In all cases, we repeated training for each set of parameters five times, each with a different random seed set in Python’s numpy, tensorflow, and random packages. All models trained for a given evaluation session and seed combination were evaluated on the same set of sentences from that evaluation session, which were held out from training and validation sets. In Fig. 4 and Supplementary Fig. 1, all traces represent the mean PER across seeds, and shaded regions represent the standard error of the mean across seeds.

For the first sweep presented in Fig. 4A and Supplementary Fig. 1A, we trained models using just one session’s data, and swept the number of sentences from that session that were used to train the phoneme decoder. Sentence counts swept were inclusively between 20 and 160 at 20 sentence increments. For sessions that had fewer than 180 total sentences (160 training sentences, 10 validation sentences, 10 evaluation sentences), an additional model was trained for five random seeds that used all available sentences from the session (all remaining sentences after removing the 20 sentences for validation and evaluation).

For the second sweep presented in Fig. 4B and D and Supplementary Fig. 1B and D, we trained models by sweeping across two parameters: the number of sessions included in the training set and the number of sentences from the evaluation session on which the model was finetuned. We selected a number of sessions from 1, 5, 10, 15, and 20 for T16 or 1, 5, 10 and 15 for T22, and a number of finetuning sentences from 0, 1, 5, 10, 30, and 50 for both participants. To control for inter-session variability, we finetuned and evaluated all models on the same final evaluation session, and the number of sessions included in the training set were counted backwards from the evaluation session. All models were trained with 50 sentences from each session included in the pretraining set of sessions, and only sessions with at least 70 sentences were included (50 training, 10 validation, 10 evaluation). Models with 0 finetuning sentences were not trained further after the initial pretraining stage, but were still evaluated on the same set of evaluation sentences as the other models after finetuning. The number of sessions along the horizontal axis in Fig. 4B and Supplementary Fig. 1B represents the number of pretraining sessions plus one for the finetuning and evaluation session.

For the third sweep presented in Fig. 4C and Supplementary Fig. 1C, for each evaluation session, we first pretrained a model on all previous sessions. Then, we finetuned the pretrained model on sentences from the evaluation session. Finally, we evaluated model performance on heldout sentences from the evaluation session as well as sentences from future sessions without any further finetuning. All sentences from all sessions were split into evaluation and validation sets of 10 sentences each, and the remaining sentences from each session were used for training. We evaluated just the evaluation sets of sentences from future sessions.

To estimate training data recommendations to achieve certain performance thresholds, we fit exponential curves (equation 1) to all data points across all evaluation sessions and computed the independent variable value (i.e. number of training sentences, number of sessions, etc…) corresponding to a specified PER.

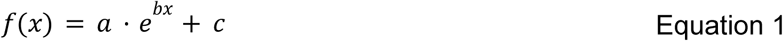

For example, to estimate the relationship between the number of training sentences from a single session and PER (in Fig. 4A), we combined all training sentence counts into a one-dimensional independent variable array (x), combined the PERs after averaging across seeds into a one-dimensional dependent variable array (f(x); equal in length to x), and fit an exponential curve between those arrays using SciPy’s curve_fit function. To estimate the independent variable (e.g., number of training sentences) corresponding to a desired PER, we evaluated the inverse of equation 1. To obtain a standard deviation around the estimated independent variable, we used the delta method ^30^:

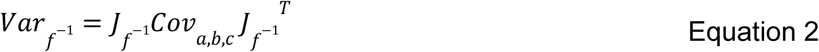

where standard deviation presented in the results is computed as Varf-1; Jf-1 is the Jacobian of the inverse of equation 1 over parameters a, b, and c; and Cova,b,c is the covariance matrix for the parameters fit by SciPy’s curve_fit function. PERs were selected within the range of data used to fit equation 1 to ensure accuracy of the delta method.

## Supplementary Tables

**Supplementary Table 1:**
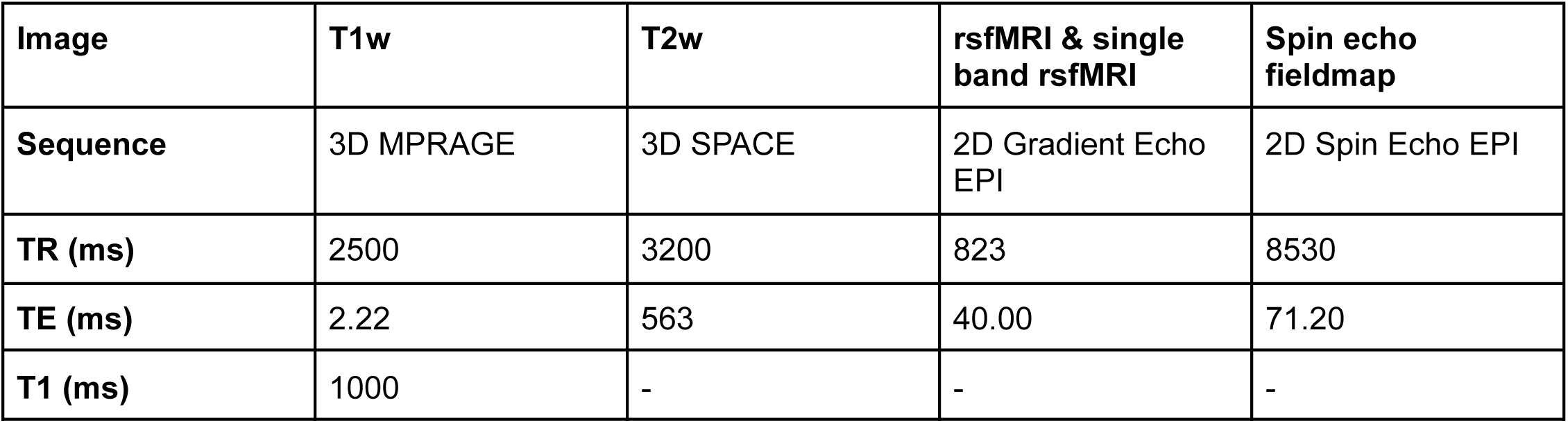
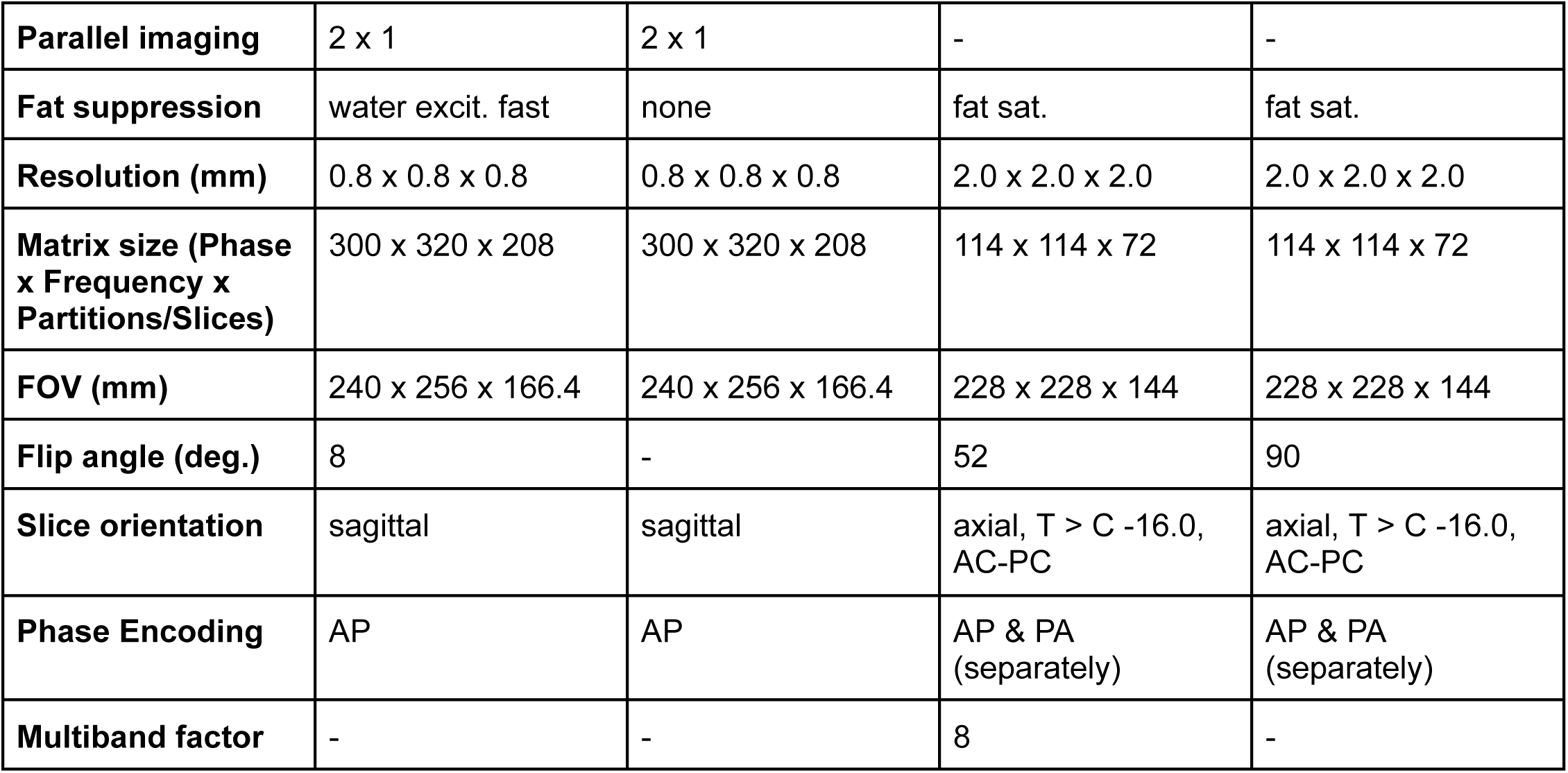
T16 MRI scan parameters.

**Supplementary Table 2:** T16 speech data collection sessions.

| Session number | Session date | Post-implant day | Data description |
|---|---|---|---|
| 1 | 01-2024<br>Session 1 | 32 | • Imagery: vocal, fast<br>• 197 offline Switchboard sentences |
| 2 | 01-2024<br>Session 2 | 36 | • Imagery: vocal, fast<br>• 177 offline Switchboard sentences |
| 3 | 01-2024<br>Session 3 | 50 | • Imagery: vocal, fast<br>• 183 offline Switchboard sentences |
| 4 | 02-2024<br>Session 1 | 64 | • Imagery: vocal and mimed, fast<br>• 381 offline 50 vocab sentences |
| 5 | 02-2024<br>Session 2 | 76 | • Imagery: mimed, fast<br>• 186 offline Switchboard sentences |
| 6 | 03-2024<br>Session 1 | 95 | • Imagery: mimed, fast<br>• 98 offline Switchboard sentences<br>• 96 evaluation Switchboard sentences |
| 7 | 03-2024<br>Session 2 | 99 | • Imagery: mimed, fast<br>• 113 offline 50 vocab sentences |
| 8 | 03-2024<br>Session 3 | 106 | • Imagery: mimed, fast<br>• 148 offline 50 vocab sentences<br>• 49 online finetuning 50 vocab sentences (finetuned with 8 sessions)<br>• 61 evaluation 50 vocab sentences |
| 9 | 04-2024<br>Session 1 | 120 | • Imagery: mimed, fast<br>• 331 offline Switchboard sentences |
| 10 | 04-2024<br>Session 2 | 134 | • Imagery: mimed, fast<br>• 244 offline Switchboard sentences<br>• 50 online finetuning Switchboard sentences (finetuned with 11 sessions)<br>• 49 evaluation Switchboard sentences |
| 11 | 04-2024<br>Session 3 | 141 | • Imagery: mimed, fast<br>• 198 offline Switchboard sentences<br>• 49 online finetuning Switchboard sentences (finetuned with 12 sessions)<br>• 25 evaluation Switchboard sentences |
| 12 | 05-2024<br>Session 1 | 148 | • Imagery: mimed, fast<br>• 263 offline Switchboard sentences<br>• 50 online finetuning Switchboard sentences (finetuned with 8 sessions)<br>• 50 evaluation Switchboard sentences |
| 13 | 05-2024<br>Session 2 | 155 | • Imagery: mimed, fast<br>• 247 offline Switchboard sentences<br>• 50 online finetuning Switchboard sentences (finetuned with 8 sessions)<br>• 25 evaluation Switchboard sentences |
| 14 | 06-2024<br>Session 1 | 204 | • Imagery: mimed, fast<br>• 246 offline Switchboard sentences<br>• 50 online finetuning Switchboard sentences (finetuned with 15 sessions)<br>• 25 evaluation Switchboard sentences |
| 15 | 07-2024<br>Session 1 | 218 | • Imagery: mimed, fast<br>• 243 offline Switchboard sentences |
| 16 | 07-2024<br>Session 2 | 237 | • Imagery: mimed, fast<br>• 93 offline Switchboard sentences |
| 17 | 08-2024<br>Session 1 | 256 | • Imagery: mimed, fast<br>• 199 offline Switchboard sentences |
| 18 | 09-2024<br>Session 1 | 277 | • Imagery: mimed, fast<br>• 97 offline Switchboard sentences |
| 19 | 09-2024<br>Session 2 | 284 | • Imagery: mimed, fast<br>• 158 diagnostic trials<br>• 144 online finetuning Switchboard sentences |
| 20 | 09-2024<br>Session 3 | 286 | • Imagery: mimed, fast<br>• 99 offline Switchboard sentences |
| 21 | 09-2024<br>Session 4 | 291 | • Imagery: mimed, fast<br>• 196 offline Switchboard sentences |
| 22 | 09-2024<br>Session 5 | 293 | • Imagery: mimed, fast<br>• 80 diagnostic trials |
| 23 | 10-2024<br>Session 1 | 314 | • Imagery: mimed, fast<br>• 158 diagnostic trials<br>• 247 offline Switchboard sentences |
| 24 | 10-2024<br>Session 2 | 319 | • Imagery: mimed, fast<br>• 49 online finetuning Switchboard sentences |
| 25 | 10-2024<br>Session 3 | 321 | • Imagery: mimed, fast<br>• 99 online finetuning Switchboard sentences |
| 26 | 10-2024<br>Session 4 | 323 | • Imagery: mimed, fast<br>• 80 diagnostic trials<br>• 90 online finetuning Switchboard sentences |
| 27 | 11-2024<br>Session 1 | 330 | • Imagery: mimed, fast<br>• 76 diagnostic trials<br>• 93 online finetuning Switchboard sentences |
| 28 | 11-2024<br>Session 2 | 333 | • Imagery: mimed, fast<br>• 80 diagnostic trials<br>• 94 online finetuning Switchboard sentences |
| 29 | 11-2024<br>Session 3 | 335 | • Imagery: mimed, fast<br>• 96 online finetuning Switchboard sentences |
| 30 | 11-2024<br>Session 4 | 340 | • Imagery: mimed, fast<br>• 92 online finetuning Switchboard sentences |
| 31 | 11-2024<br>Session 5 | 344 | • Imagery: mimed, fast<br>• 79 diagnostic trials<br>• 78 online finetuning Switchboard sentences (finetuned with 10 sessions)<br>• 79 evaluation Switchboard sentences |
| 32 | 11-2024<br>Session 6 | 347 | • Imagery: mimed, slow<br>• 80 diagnostic trials<br>• 118 online finetuning Switchboard sentences (finetuned with 10 sessions)<br>• 80 evaluation Switchboard sentences<br>• 100 offline 50 vocab sentences |
| 33 | 11-2024<br>Session 7 | 349 | • Imagery: mimed, slow<br>• 47 online finetuning Switchboard sentences |
| 34 | 11-2024<br>Session 8 | 351 | • Imagery: mimed, slow<br>• 40 online finetuning Switchboard sentences |
| 35 | 11-2024<br>Session 9 | 356 | • Imagery: mimed, slow<br>• 80 diagnostic trials<br>• 77 online finetuning Switchboard sentences (finetuned with 10 sessions)<br>• 115 evaluation Switchboard sentences<br>• 50 online finetuning 50 vocab sentences (finetuned with 3 sessions)<br>• 40 evaluation 50 vocab sentences |
| 36 | 12-2024<br>Session 1 | 361 | • Imagery: mimed, slow<br>• 80 diagnostic trials<br>• 40 online finetuning Switchboard sentences |
| 37 | 12-2024<br>Session 2 | 363 | • Imagery: mimed, slow<br>• 40 online finetuning Switchboard sentences |
| 38 | 12-2024<br>Session 3 | 365 | • Imagery: mimed, slow<br>• 49 online finetuning Switchboard sentences |
| 39 | 12-2024<br>Session 4 | 370 | • Imagery: mimed, slow<br>• 79 diagnostic trials<br>• 50 online finetuning Switchboard sentences |
| 40 | 01-2025<br>Session 1 | 403 | • Imagery: mimed, slow<br>• 55 online finetuning Switchboard sentences |
| 41 | 01-2025<br>Session 2 | 419 | • Imagery: mimed, slow<br>• 73 online finetuning Switchboard sentences |
| 42 | 02-2025<br>Session 1 | 428 | • Imagery: mimed, slow<br>• 60 online finetuning Switchboard sentences (finetuned with 10 sessions)<br>• 19 evaluation Switchboard sentences |
| 43 | 02-2025<br>Session 2 | 431 | • Imagery: mimed, slow<br>• 60 online finetuning Switchboard sentences |
| 44 | 02-2025<br>Session 3 | 433 | • Imagery: mimed, slow<br>• 74 online finetuning Switchboard sentences |
| 45 | 02-2025<br>Session 4 | 435 | • Imagery: mimed, slow<br>• 80 diagnostic trials<br>• 40 online finetuning Switchboard sentences |
| 46 | 02-2025<br>Session 5 | 445 | • Imagery: mimed, slow<br>• 75 diagnostic trials<br>• 66 online finetuning Switchboard sentences |
| 47 | 02-2025<br>Session 6 | 447 | • Imagery: mimed, slow<br>• 97 online finetuning Switchboard sentences |
| 48 | 03-2025<br>Session 1 | 452 | • Imagery: mimed, slow<br>• 80 diagnostic trials<br>• 49 online finetuning Switchboard sentences |
| 49 | 03-2025<br>Session 2 | 459 | • Imagery: mimed, slow<br>• 80 diagnostic trials<br>• 71 online finetuning Switchboard sentences |
| 50 | 03-2025<br>Session 3 | 473 | • Imagery: mimed, slow<br>• 80 diagnostic trials<br>• 58 online finetuning Switchboard sentences (finetuned with 20 sessions)<br>• 20 evaluation Switchboard sentences |
| 51 | 03-2025<br>Session 4 | 475 | • Imagery: mimed, slow<br>• 72 online finetuning Switchboard sentences |
| 52 | 04-2025<br>Session 1 | 487 | • Imagery: mimed, slow<br>• 80 diagnostic trials<br>• 71 online finetuning Switchboard sentences (finetuned with 20 sessions)<br>• 20 evaluation Switchboard sentences |
| 53 | 04-2025<br>Session 2 | 491 | • Imagery: mimed, slow<br>• 36 online finetuning Switchboard sentences |
| 54 | 04-2025<br>Session 3 | 494 | • Imagery: mimed, slow<br>• 91 online finetuning Switchboard sentences |
| 55 | 04-2025<br>Session 4 | 501 | • Imagery: mimed, slow<br>• 80 diagnostic trials<br>• 92 online finetuning Switchboard sentences (finetuned with 20 sessions)<br>• 29 evaluation Switchboard sentences |
| 56 | 04-2025<br>Session 5 | 510 | • Imagery: mimed, slow<br>• 79 diagnostic trials<br>• 80 online finetuning Switchboard sentences (finetuned with 20 sessions)<br>• 38 evaluation Switchboard sentences |
| 57 | 05-2025<br>Session 1 | 515 | • Imagery: mimed, slow<br>• 80 diagnostic trials<br>• 71 online finetuning Switchboard sentences (finetuned with 20 sessions)<br>• 51 evaluation Switchboard sentences |
| 58 | 05-2025<br>Session 2 | 526 | • Imagery: mimed, slow<br>• 77 diagnostic trials<br>• 44 online finetuning Switchboard sentences (finetuned with 20 sessions)<br>• 48 evaluation Switchboard sentences |
| 59 | 05-2025<br>Session 3 | 538 | • Imagery: mimed, slow<br>• 80 diagnostic trials<br>• 51 online finetuning Switchboard sentences |
| 60 | 06-2025<br>Session 1 | 545 | • Imagery: mimed, slow<br>• 79 diagnostic trials<br>• 60 online finetuning Switchboard sentences |
| 61 | 06-2025<br>Session 2 | 547 | • Imagery: mimed, slow<br>• 79 diagnostic trials<br>• 70 online finetuning Switchboard sentences (finetuned with 10 sessions)<br>• 40 evaluation Switchboard sentences |
| 62 | 06-2025<br>Session 3 | 550 | • Imagery: mimed, slow<br>• 39 online finetuning Switchboard sentences |
| 63 | 06-2025<br>Session 4 | 554 | • Imagery: mimed, slow<br>• 79 diagnostic trials<br>• 72 online finetuning Switchboard sentences (finetuned with 20 sessions)<br>• 29 evaluation Switchboard sentences |
| 64 | 06-2025<br>Session 5 | 557 | • Imagery: mimed, slow<br>• 39 online finetuning Switchboard sentences |
| 65 | 06-2025<br>Session 6 | 561 | • Imagery: mimed, slow<br>• 158 diagnostic trials<br>• 109 online finetuning Switchboard sentences (finetuned with 20 sessions)<br>• 70 evaluation Switchboard sentences |
| 66 | 07-2025<br>Session 1 | 573 | • Imagery: mimed, slow<br>• 77 diagnostic trials<br>• 60 online finetuning Switchboard sentences |
| 67 | 07-2025<br>Session 2 | 580 | • Imagery: mimed, slow<br>• 60 online finetuning Switchboard sentences |
| 68 | 07-2025<br>Session 3 | 594 | • Imagery: mimed, slow<br>• 59 online finetuning Switchboard sentences |
| 69 | 07-2025<br>Session 4 | 596 | • Imagery: mimed, slow<br>• 79 diagnostic trials<br>• 114 online finetuning Switchboard sentences (finetuned with 20 sessions)<br>• 40 evaluation Switchboard sentences |
| 70 | 08-2025<br>Session 1 | 603 | • Imagery: mimed, slow<br>• 80 diagnostic trials<br>• 112 online finetuning Switchboard sentences (finetuned with 10 sessions)<br>• 79 evaluation Switchboard sentences |
| 71 | 09-2025<br>Session 1 | 638 | • Imagery: mimed, slow<br>• 79 diagnostic trials<br>• 122 online finetuning Switchboard sentences (finetuned with 20 sessions)<br>• 37 evaluation Switchboard sentences |
| 72 | 09-2025<br>Session 2 | 659 | • Imagery: mimed, slow<br>• 80 diagnostic trials<br>• 158 online finetuning Switchboard sentences (finetuned with 20 sessions)<br>• 35 evaluation Switchboard sentences |
| 73 | 09-2025<br>Session 3 | 662 | • Imagery: mimed, slow<br>• 80 diagnostic trials<br>• 117 online finetuning Switchboard sentences (finetuned with 20 sessions)<br>• 36 evaluation Switchboard sentences |
| 74 | 10-2025<br>Session 1 | 673 | • Imagery: mimed, slow<br>• 79 diagnostic trials |
| 75 | 10-2025<br>Session 2 | 680 | • Imagery: mimed, slow<br>• 80 diagnostic trials |
| 76 | 10-2025<br>Session 3 | 687 | • Imagery: mimed, slow<br>• 80 diagnostic trials<br>• 156 online finetuning Switchboard sentences |
| 77 | 10-2025<br>Session 4 | 692 | • Imagery: mimed, slow<br>• 59 online finetuning Switchboard sentences |
| 78 | 11-2025<br>Session 1 | 697 | • Imagery: mimed, slow<br>• 94 online finetuning Switchboard sentences |
| 79 | 11-2025<br>Session 2 | 701 | • Imagery: mimed, slow<br>• 78 online finetuning Switchboard sentences |
| 80 | 11-2025<br>Session 3 | 704 | • Imagery: mimed, slow<br>• 80 diagnostic trials<br>• 116 online finetuning Switchboard sentences (finetuned with 20 sessions)<br>• 40 evaluation Switchboard sentences |
| 81 | 11-2025<br>Session 4 | 708 | • Imagery: mimed, slow<br>• 45 online finetuning Switchboard sentences |
| 82 | 11-2025<br>Session 5 | 718 | • Imagery: mimed, slow<br>• 43 online finetuning Switchboard sentences |
| 83 | 12-2025<br>Session 1 | 736 | • Imagery: mimed, slow<br>• 80 diagnostic trials<br>• 76 online finetuning Switchboard sentences (finetuned with 10 sessions)<br>• 39 evaluation Switchboard sentences |

**Supplementary Table 3:** T22 speech data collection sessions.

| Session number | Session date | Post-implant day | Data description |
|---|---|---|---|
| 1 | 2026-03<br>Session 1 | 19 | • Imagery: mimed<br>• 170 training OpenSubtitles sentences<br>• 30 evaluation OpenSubtitles sentences |
| 2 | 2026-03<br>Session 2 | 20 | • Imagery: mimed<br>• 166 training OpenSubtitles sentences<br>• 30 evaluation OpenSubtitles sentences |
| 3 | 2026-03<br>Session 3 | 21 | • Imagery: mimed<br>• 217 training OpenSubtitles sentences<br>• 30 evaluation OpenSubtitles sentences |
| 4 | 2026-03<br>Session 4 | 26 | • Imagery: mimed<br>• 131 training OpenSubtitles sentences<br>• 30 evaluation OpenSubtitles sentences |
| 5 | 2026-03<br>Session 5 | 27 | • Imagery: mimed<br>• 289 training OpenSubtitles sentences<br>• 30 evaluation OpenSubtitles sentences |
| 6 | 2026-04<br>Session 1 | 28 | • Imagery: mimed<br>• 327 training OpenSubtitles sentences<br>• 30 evaluation OpenSubtitles sentences |
| 7 | 2026-04<br>Session 2 | 33 | • Imagery: mimed<br>• 220 training OpenSubtitles sentences<br>• 30 evaluation OpenSubtitles sentences |
| 8 | 2026-04<br>Session 3 | 34 | • Imagery: mimed<br>• 184 training OpenSubtitles sentences<br>• 30 evaluation OpenSubtitles sentences |
| 9 | 2026-04<br>Session 4 | 35 | • Imagery: mimed<br>• 180 training OpenSubtitles sentences<br>• 30 evaluation OpenSubtitles sentences |
| 10 | 2026-04<br>Session 5 | 43 | • Imagery: mimed<br>• 86 training OpenSubtitles sentences<br>• 30 evaluation OpenSubtitles sentences |
| 11 | 2026-04<br>Session 6 | 44 | • Imagery: mimed<br>• 138 training OpenSubtitles sentences<br>• 30 evaluation OpenSubtitles sentences |
| 12 | 2026-04<br>Session 7 | 48 | • Imagery: mimed<br>• 477 training Switchboard sentences<br>• 30 evaluation Switchboard sentences |
| 13 | 2026-04<br>Session 8 | 56 | • Imagery: mimed<br>• 292 training Switchboard sentences<br>• 30 evaluation Switchboard sentences |
| 14 | 2026-04<br>Session 9 | 57 | • Imagery: mimed<br>• 315 training Switchboard sentences<br>• 30 evaluation Switchboard sentences |
| 15 | 2026-05<br>Session 1 | 63 | • Imagery: mimed<br>• 187 training Switchboard sentences<br>• 30 evaluation Switchboard sentences |
| 16 | 2026-05<br>Session 2 | 64 | • Imagery: mimed<br>• 240 training Switchboard sentences<br>• 30 evaluation Switchboard sentences |
| 17 | 2026-05<br>Session 3 | 70 | • Imagery: mimed<br>• 223 training Switchboard sentences<br>• 30 evaluation Switchboard sentences |
| 18 | 2026-05<br>Session 4 | 71 | • Imagery: mimed<br>• 126 training Switchboard sentences<br>• 30 evaluation Switchboard sentences |

**Supplementary Table 4:** Software packages.

| **Software** | **Version** | **Description** |
|---|---|---|
| Python | 3.9 | Used for realtime brain-to-text decoding and all analysis |
| TensorFlow | 2.10 | Used for phoneme decoders |
| SciPy | 1.12 | Used to fit exponential curves to data recommendation sweeps |
| G2P_En | 2.1 | Used to convert sentences into phoneme sequences |
| Ubuntu | 22.04 | Used in session on all data collection machines, except one Windows computer |
| Windows | 10 v1809 | Used in session for raw neural activity recording and storage to disk |

**Supplementary Table 5:** Decoder hyperparameters.

| Parameter category | Parameter name | Parameter description | T16 parameter value | T22 parameter value |
|---|---|---|---|---|
| Offline phoneme decoder | RNN layers | Number of GRU layers | 5 | 5 |
|  | RNN units | Number of units per GRU layer | 512 | 512 |
|  | RNN dropout | Probability of unit dropout in the GRU layers | 0.4 | 0.4 |
|  | RNN kernel size | Number of input feature time bins stacked as a single input for the RNN | 14 | 14 |
|  | RNN kernel stride | Number of time steps the RNN skips forward each step | 4 | 4 |
|  | Input units | Number of units in each input layer | 512 | 512 |
|  | Input dropout | Probability of unit dropout in the input layers | 0.2 | 0.2 |
|  | Input activation | Activation function for input layer units | softsign | softsign |
|  | Augmentation white noise SD | Standard deviation of white noise added to input data during training | 0.5 | 0.5 |
|  | Augmentation smoothing kernel SD | Standard deviation of the Gaussian kernel used to smooth input data during training | 2 | 2 |
|  | Augmentation constant offset SD | Standard deviation of constant offset noise added to input data during training | 0.2 | 0.2 |
|  | Batch size | Number of sentences included in each batch | 64 | 64 |
|  | Training batches | Maximum number of training batches | 10,000 | 10,000 |
|  | Early stop batches | Number of training batches without a validation PER improvement to stop training early | 3,000 | 3,000 |
|  | Starting learning rate | Initial learning rate at the start of training | 0.01 | 0.01 |
|  | Learning rate decay | The scalar by which learning rate decayed | 0.9 | 0.9 |
|  | Learning rate patience | The number of batches that must have passed without improvement in validation loss to reduce learning rate | 300 | 300 |
|  | *β*1 | ADAM stochastic gradient descent parameter | 0.9 | 0.9 |
|  | *β*2 | ADAM stochastic gradient descent parameter | 0.999 | 0.999 |
|  | *ϵ* | ADAM stochastic gradient descent parameter | 0.1 | 0.1 |
| Finetuned phoneme decoder (same as offline except where noted) | New data percent | Percent of sentences per batch from the new session | 0.6 | 0.6 |
| Online phoneme decoder (same as finetuned except where noted) | Constant learning rate | Learning rate used during online fine-tuning | 0.004 | 0.004 |
|  | Min training steps | Minimum number of training epochs to execute | 32 | 32 |
|  | Max training steps | Maximum number of training epochs to execute | 200 | 200 |
|  | Loss threshold | Loss threshold to stop training once under | 0.5 | 0.5 |
| 50-word language model | 5-gram blank penalty | Penalty applied to blank tokens | log(2) | - |
|  | 5-gram acoustic scale | Scaling factor on RNN’s log probabilities | 0.8 | - |
|  | 5-gram beam min active | Beam search decoder’s minimum active states | 200 | - |
|  | 5-gram beam max active | Beam search decoder’s maximum active states | 7,000 | - |
|  | 5-gram beam size | Size of the beam | 17 | - |
|  | 5-gram number best | Number of decoding hypotheses | 1 | - |
| 1,024-word language model (same as 50-word except where noted) | 5-gram blank penalty | Penalty applied to blank tokens | log(4) | log(4) |
|  | 5-gram acoustic scale | Scaling factor on RNN’s log probabilities | 0.325 | 0.35 |
|  | 5-gram number best | Number of decoding hypotheses | 100 | 100 |
|  | Alpha | Interpolation factor between the 5-gram and OPT LMs | 0.65 | 0.45 |
| 125k-word language model (same as 1,024-word except where noted) | 5-gram blank penalty | Penalty applied to blank tokens | log(6) | log(6) |
|  | 5-gram acoustic scale | Scaling factor on RNN’s log probabilities | 0.25 | 0.35 |
|  | Alpha | Interpolation factor between the 5-gram and OPT LMs | 0.7 | 0.475 |

**Supplementary Table 6:** Results summary.

| Category | Metric | T16 | T22 | Prior work |
|---|---|---|---|---|
| 125,000-word vocabulary | PER, RNN output (%) | 21.80 [18.54–27.14] | 14.43 [12.82–18.40] | 20.9 (11), 16.7 (12) |
| 125,000-word vocabulary | PER after language model (%) | 12.28 [8.84–17.70] | 5.83 [4.63–10.40] | — |
| 125,000-word vocabulary | WER after language model (%) | 19.37 [13.83–26.50] | 10.62 [7.57–15.67] | 24.7 (11), 9.22 (12) |
| 125,000-word vocabulary | Speaking rate (WPM) | 34.98 [29.83–43.39] | 35.31 [33.38–36.38] | 63 (11), 52.3 (12) |
| 1,024-word vocabulary | PER, RNN output (%) | 16.11 [14.47–22.39] | 7.43 [6.22–10.53] | 29.4 (10) |
| 1,024-word vocabulary | PER after language model (%) | 6.17 [2.44–7.14] | 1.18 [0.69–3.44] | — |
| 1,024-word vocabulary | WER after language model (%) | 10.00 [4.35–13.63] | 2.25 [0.93–5.32] | 25.5 (10) |
| 1,024-word vocabulary | Speaking rate (WPM) | 34.88 [31.14–48.16] | 36.54 [35.18–38.71] | 78.3 (10) |
| 50-word vocabulary | PER, RNN output (%) | 14.31 [10.88–16.86] | — | 22.1 (11), 4.93 (12) |
| 50-word vocabulary | PER after language model (%) | 9.13 [5.94–11.77] | — | — |
| 50-word vocabulary | WER after language model (%) | 11.89 [8.88–13.49] | — | 25.6 (9), 11.2 (11), 0.22 (12) |
| 50-word vocabulary | Speaking rate (WPM) | 52.47 [47.69–54.77] | — | 15.2 (9), 58 (11), 39.11 (12) |
| Q&A (spontaneous responses) | PER, RNN output (%) | 33.65 [30.76–34.97] | — | — |
| Q&A (spontaneous responses) | PER after language model (%) | 22.62 [18.83–26.55] | — | — |
| Q&A (spontaneous responses) | WER after language model (%) | 35.21 [28.00–38.68] | — | — |
| Q&A (spontaneous responses) | Speaking rate (WPM) | 27.74 [25.64–29.62] | — | — |
| Single-session calibration | Estimated minimum PER, curve plateau (%) | 32.26 ± 1.59 | 27.09 ± 0.87 | — |
| Single-session calibration | Sentences to reach plateau + 5% of fitted range | 122.9 ± 13.1 | 115.7 ± 4.5 | — |
| Single-session calibration | Recording time for those sentences (min) | 21.43 ± 2.28 | 19.98 ± 0.78 | — |
| Multi-session pretraining | Estimated minimum PER, curve plateau (%) | 27.96 ± 1.55 | 28.45 ± 0.93 | — |
| Multi-session pretraining | Sessions (50 sentences each) to reach plateau + 5% of fitted range | 13.0 ± 3.6 | 7.0 ± 1.3 | — |
| Decoder drift without recalibration | Plateau of PER relative to same-day PER (×) | 2.10 ± 0.24 | Not estimable (trend not exponential) | — |
| Decoder drift without recalibration | Days until PER rises 25% (1.25×) | 6.9 ± 1.8 | Not estimable | — |
| Fine-tuning in a new session | Plateau of PER relative to no fine-tuning (×) | 0.69 ± 0.04 | 0.59 ± 0.19 | — |
| Fine-tuning in a new session | Sentences to reach 0.75× (25% PER reduction) | 35.5 ± 4.3 | 56.9 ± 7.3 | — |
| Fine-tuning in a new session | Share of possible reduction reached at 0.75× (%) | 81.2 | 61.3 | — |
| Fine-tuning in a new session | Recording time to reach 0.75× (min) | 6.19 ± 0.75 | 9.83 ± 1.26 | — |
| Fine-tuning in a new session | Sentences to reach 0.80× (20% PER reduction) | 22.4 ± 2.7 | 40.7 ± 2.8 | — |
| Fine-tuning in a new session | Share of possible reduction reached at 0.80× (%) | 65.1 | 49.3 | — |
| Fine-tuning in a new session | Recording time to reach 0.80× (min) | 3.90 ± 0.47 | 7.03 ± 0.48 | — |
| Recording channels | Channels modulated during speech (mean ± SD across sessions) | 56 ± 3.1 of 64 | 242 ± 8.8 of 256 | — |

## Supplementary figures

**Supplementary Fig. 1:**
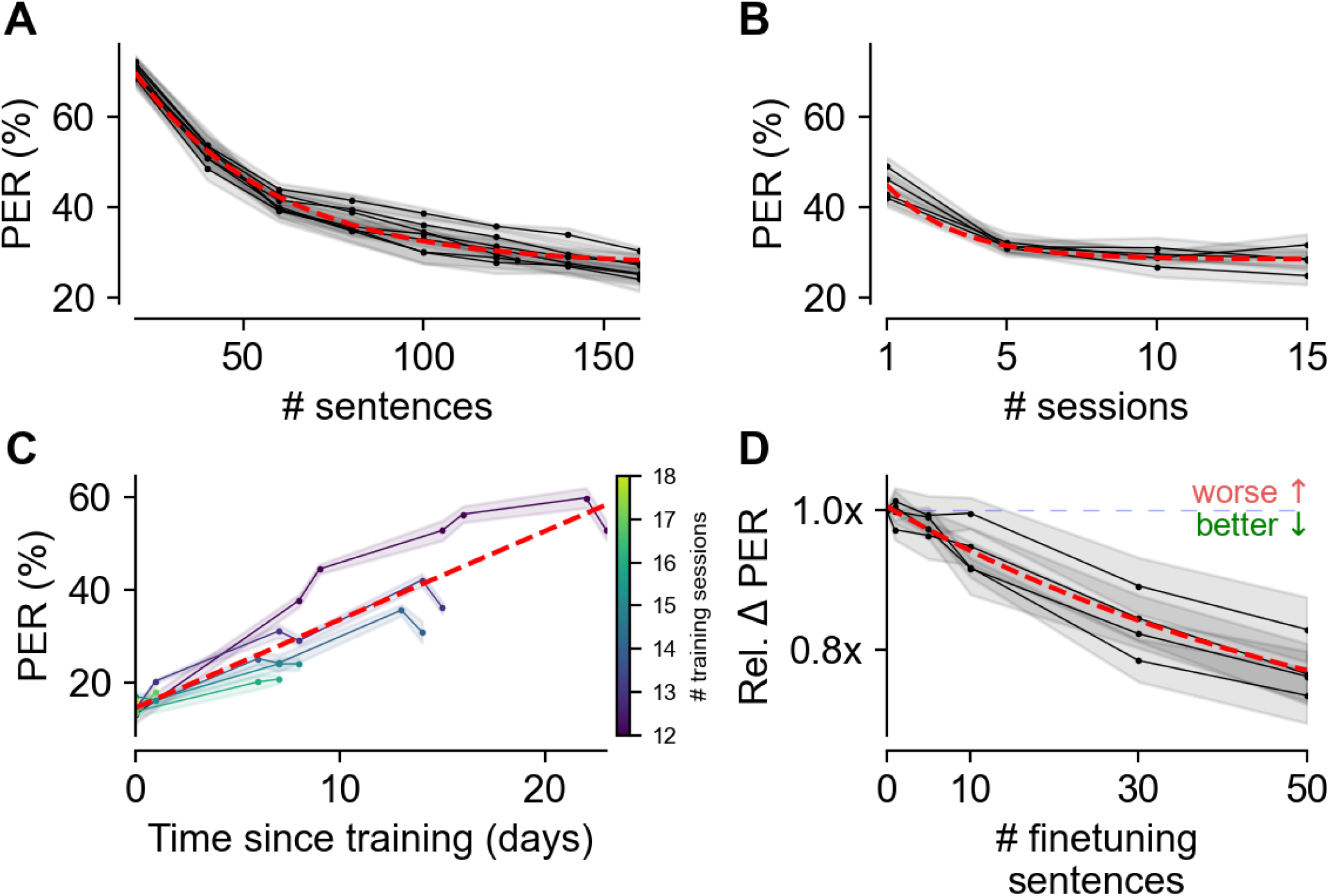
Data scaling properties for brain-to-text decoding for participant T22. (**A**) PERs resulting from training a phoneme decoder on a select number of calibration sentences from a single T22 session. Seven sessions are shown. The red dashed trace represents an exponential fit to all data points. Dots represent evaluation points. (**B**) PERs resulting from varying the number of calibration sessions with 50 calibration sentences from each T22 session. (**C**) PER from frozen models evaluated on sentences from future T22 sessions. The models trained on more sessions are more stable on future sessions. (**D**) Relative change in PER resulting from finetuning a previously trained model on a small set of calibration sentences from a new T22 session. Relative change in PER is computed as the ratio between the PER after finetuning on the indicated number of sentences relative to the PER without any finetuning. In all panels, traces represent the mean PER across five random seeds and shaded regions represent the standard error of the mean across seeds.

